# Use of Recommended Real-World Methods for Electronic Health Record Data Analysis Has Not Improved Over 10 Years

**DOI:** 10.1101/2023.06.21.23291706

**Authors:** Chenyu Li, Abdulrahman M. Alsheikh, Karen A. Robinson, Harold P. Lehmann

## Abstract

**Background and Purpose:** To document the use of recommended Real-World Methods (RWM) in Electronic Health Record (EHR)-based analysis in biomedical research over 10 years.

**Methods:** Sampled-article scoping review of methods used in EHR-based biomedical research. We developed a search strategy to identify reports of biomedical research based on EHR data and systematically sampled articles from different ranges of years (epochs) between 2010 and 2019 to establish a trajectory of use of recommended RWM. Methods were classified by 3 phases of research: pre-analytic (missing data), analytic (specific methods), and post-analytic (sensitivity analysis). The primary outcome was the proportion of studies using recommended RWM within each epoch. Meta-regressions were performed to examine trends.

**Data Synthesis:** Five epochs were defined between 2010 and 2019 with 35 studies selected per epoch as pre-defined by a sample size calculation. Of the 175 articles reviewed, 70 (40.%) reported recommended RWM in any of the 3 phases of research. The breakdown for the most recent year in the dataset, 2019, was 14.% (95% confidence interval 2.7%, 26.%), 14.% (2.7%, 26.%), and 11.% (0.89%, 22.%), for assessing missing data, using specific methods, and performing sensitivity analysis, respectively. Only 3.4 % of studies used appropriate methods for each phase of research. Meta-regression slopes for each of the three phases were statistically 0.

**Limitation and Conclusions:** The underuse of recommended Real-World Methods (RWM) in EHR-based biomedical research remains a concern, with less than 50% of reports using these methods in any phase of research over the last decade. This lack of use indicates a continued risk of bias in the EHR-based literature.

## INTRODUCTION

Use of Real-World Data (RWD) to generate Real-World Evidence (RWE) is playing an increasing role in health care decisions worldwide.[1] RWD refers to information acquired from a range of sources generated in the process of clinical care.[1] Electronic health records (EHRs), claims databases, patient registries, and other types of data repositories may be among these sources.[1,2] Turning RWD into RWE can provide useful insights into the safety, effectiveness, and value of medical products and treatments in real-world clinical settings.[3] There is a growing interest in using RWE by stakeholders, including policymakers, biomedical researchers, clinicians, and medical product developers.[4–10] Despite potential broad utility, the quality of RWE, compared with evidence generated from Randomized Controlled Trials (RCTs) — still the gold-standard of clinical research[3]—is found wanting. The methods recommended to derive RWE from RWD are intended to reduce bias (especially confounding) and to allow associational and even causal inference when using observational data. Not using the appropriate methods may result in inaccurate, biased, or even inequitable algorithms.[11] Ultimately, applying such results or algorithms in real-world practice may result in the improper treatment of patients.[12]

EHRs are essential RWD sources[1]. The 21st Century Cures Act in the U.S. was signed into law in 2016 and calls on Congress to “evaluate the potential use of real-world evidence*.”*[13] FDA has developed guidelines on the various uses of RWE, for example.[2,14,15] The guidelines for data analysis and RWE generation methodology remain under discussion. [1,2,14–16] The COVID-19 pandemic has surfaced the importance of RWE for addressing new diseases,[17] while, at the same time, methodological concerns about the use of RWE continue to be raised.[18–20]

Discussions about Real World Data analysis address 3 phases: pre-analytic (missing data), analytic (specific methods), and post-analytic (sensitivity analysis). During phase 1, the pre-analytic phase (the preparation of data for analysis), addressing missing data is of special concern, because researchers have no control of what data are collected or recorded, and so traditional approaches may not apply, necessitating, for instance, greater attention to data missing not at random (MNAR).[21,22] During phase 2, the analytic phase (the statistical modeling and analysis of the data), many Analytic Real World Methods (ARWM) have been developed to analyze RWD, from the statistical, epidemiological, and computer-science communities.[19,23–25] During phase 3, the post-analytic phase, sensitivity analyses establish the robustness of conclusions to reasonable uncertainty in assumptions.[24–27] In the current paper, we call the entire set of methods in these 3 phases of research “recommended Real World Methods (recommended RWM).”

Our objective was to assess use of recommended RWM for each of the 3 phases in biomedical research assessing hypothesized associations and treatment effectiveness, based on EHR data, as documented in the peer-reviewed literature and to assess whether this use has changed over the past 10 years. We focused on this research domain, because it is the focus of much desire for using EHRs for clinical research.[28]

Because our goal was to identify proportions, we did not need to identify every relevant article published in the past 10 years as would be required for a standard systematic review. Instead, we sampled from papers identified through a thorough literature search and applied a systematic-review approach to the sample papers. We call this approach a sampled-article scoping review.

## METHODS

Our scoping review was based on randomly-sampled papers, from which we aimed (1) to define the articles that would be eligible for review (i.e., operationalize the intention of “papers that used EHR as the main data source for biomedical research”), (2) to operationalize the list of recommended RWM; and (3) to establish the proportion of use of the these methods for time epochs over the past 10 years. We followed the Preferred Reporting Items for Systematic Reviews and Meta-Analyses Extension for Scoping Review (PRISMA-ScR) checklist in reporting this scoping review. [29]

Given the novelty of our method, for items (1) and (2), we conducted a pilot review[30] to derive the search strategy, to refine our inclusion and exclusion criteria, to establish lists of keywords for identifying recommended RWM, to estimate inter-rater reliability, and to generate data needed for the sample-size calculation for the study.

### Data Sources and Searches

We sought studies that used EHR data for traditional biomedical-research aims of assessing hypothesized associations. We used an NLM-supplied search strategy for keyword “Electronic Health Records”[31] to search PubMed. We modified the NLM supplied strategy based on results from pilot; see Supplement (Appendix Table 1 Search Strategies Details) for details. We limited the publication date to 2010/01/01-2019/12/31. The search was conducted on 2020/11/09. The detailed search strategy is presented in Appendix Table 1.

We developed the list of eligibility criteria iteratively, see Table 1. Because our focus was on associational and causal studies, we excluded review papers, research reports based on data from claims, genomic analysis, manually collected registries, or prospective clinical trials. Research that was not patient focused or biomedical, such as those focused on physician behavior, information-system evaluation, health-services evaluation, and new information technology in healthcare, were also excluded. Finally, research based on unstructured and semi-structured data which needed natural language processing or text mining was excluded.

**Table 1.**
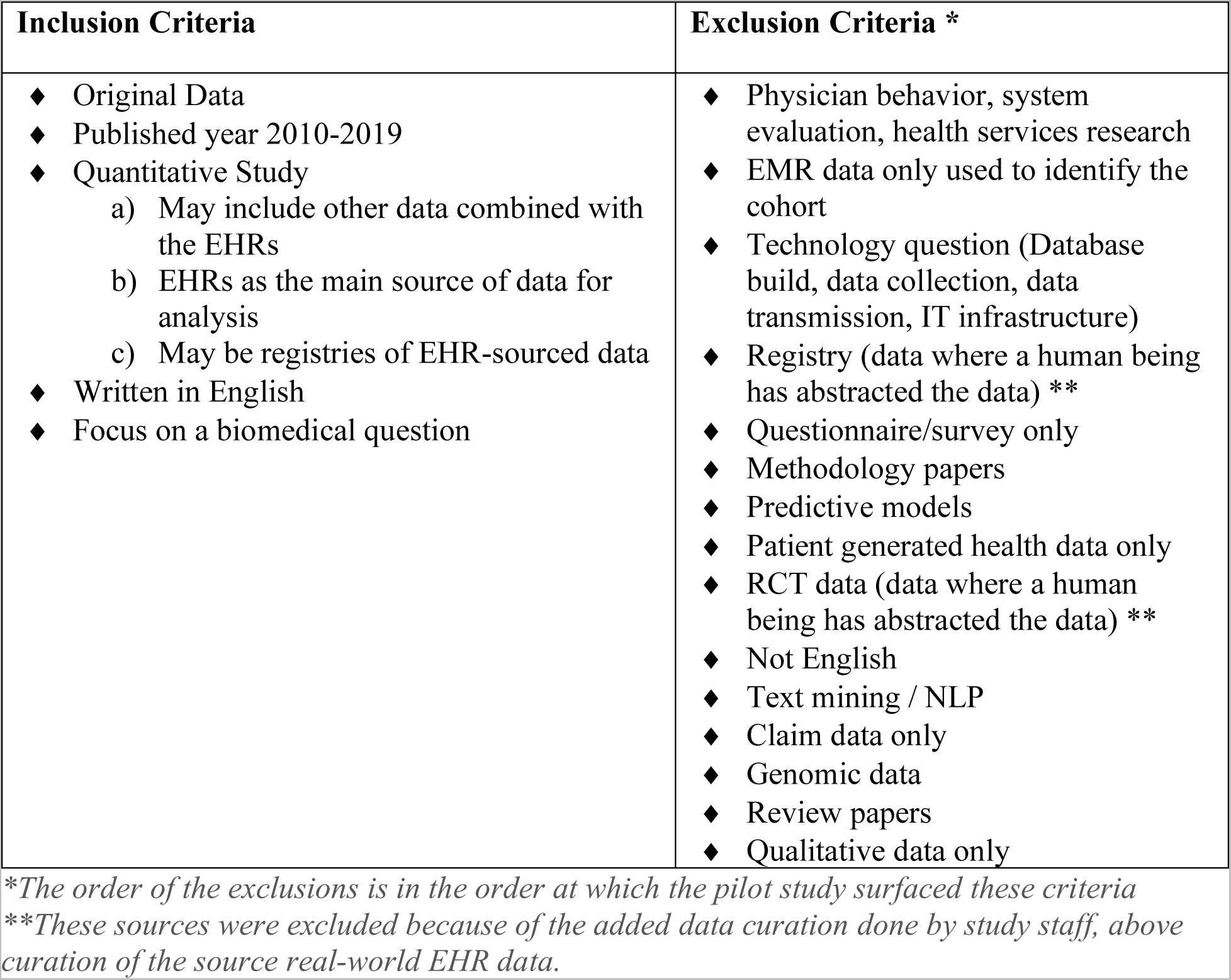
Inclusion Exclusion Criteria

| Inclusion Criteria | Exclusion Criteria * |
| --- | --- |
| ◆ Original Data<br>◆ Published year 2010-2019<br>◆ Quantitative Study<br>a) May include other data combined with the EHRs<br>b) EHRs as the main source of data for analysis<br>c) May be registries of EHR-sourced data<br>◆ Written in English<br>◆ Focus on a biomedical question | ◆ Physician behavior, system evaluation, health services research<br>◆ EMR data only used to identify the cohort<br>◆ Technology question (Database build, data collection, data transmission, IT infrastructure)<br>◆ Registry (data where a human being has abstracted the data) **<br>◆ Questionnaire/survey only<br>◆ Methodology papers<br>◆ Predictive models<br>◆ Patient generated health data only<br>◆ RCT data (data where a human being has abstracted the data) **<br>◆ Not English<br>◆ Text mining / NLP<br>◆ Claim data only<br>◆ Genomic data<br>◆ Review papers<br>◆ Qualitative data only |

*\*The order of the exclusions is in the order at which the pilot study surfaced these criteria*
*\*\*These sources were excluded because of the added data curation done by study staff, above curation of the source real-world EHR data.*

### Study Selection

We divided the 10-year period, 2010−2019 into 5 epochs: 2010−2013, 2014−2016, 2017, 2018, and 2019. We hypothesized that the proportions would rise, because of guidance that came out during the decade, specifically in the years 2013[14], 2016[32], 2018[2,15,16,33], and 2019.[34] Our goal was to get a sense of change over time, so we grouped older years together, giving more recent years finer precision. Older years had fewer articles, so we wanted our sampling to be fair to each epoch. By equalizing denominators (approximately), equal numerators made sense. We ended at 2019, as research published during the COVID-19 pandemic is likely qualitatively different.

Our statistical goal was to distinguish proportions between 2 epochs. The proportion of recommended RWM use from our pilot review was 10% (Pilot study results see Appendix Table 3, Appendix Table 4); that percentage, coupled with a desired confidence level (*α*) of 0.95, was used to calculate the sample size [35] for our scoping review of 35 eligible papers per epoch. We sampled 300 citations within each epoch, and then screened papers based on inclusion/exclusion criteria, until we reached 35 included papers per epoch. Random selection was performed by generating random numbers via Python (package *Random*) within the set of numbers representing all papers retrieved for an epoch.

Citations were downloaded and titles and abstracts were screened by one person (CL). We calculated the inter-rater reliability of CL and AA in the pilot study, the Cohen’s Kappa was used to measure inter-rater reliability, κ=0.95, and the agreement between the two reviewers was 98%. If there was any uncertainty, the article was included for full-article screening, and any remaining questions were discussed among 3 of us (HL, CL, AA).

### Data Extraction and Quality Assessment

For each randomly selected article, we recorded study type, study design and methods used. The key outcome variable was whether recommended RWM were used for each of the three phases of research. We documented the RWM methods used in included papers, then matched these to the recommended RWMs we identified from regulator guidelines, books, and RWD meeting recommendations.[2,14,15,25,26,34,36,37]

We then scored each article for whether use of a recommended RWM was reported; details are provided in Appendix Table 2. For phase 1, studies that even simply deleted records with missing data were tagged as “Addressed missing data.” Papers that reported performing any of the following were tagged as “assessed missing data”: deletion methods with examining the sensitivity of results to the missing completely at random (MCAR) and missing at random (MAR) assumptions,[26] single imputation methods, and model based methods.[26,38] For phase 2, we looked for words listed in the “Analytic Real World Method” section of Appendix Table. To broaden the definition of RWM as broadly as possible, any machine-learning methods combined with causal inference also were considered as RWM. [34] (See Appendix Table 2)

For phase 3, we looked for the words, “sensitivity” or “sensitivity analysis.”

Where judgment was needed, we biased in favor of ascribing use of RWM. Therefore, our final proportions are upper bounds within each epoch of the use of such methods. The full list of 35 RWM and 45 traditional methods is provided in Appendix Table 2.

Paper characteristics details (see Appendix Table 9 available in GitHub, due to size:^1^) were extracted by a single reader (CL), discussing ambiguous results with the senior author (HL). Beyond citation data, data collected include research design (primary objective), study design type, association type, key variables (exposure, outcome, confounding), information pertaining to the 3 phases, and use of reporting checklists (such as STROBE[39] or RECORD[40]). We also searched the supplementary materials of all the papers, looking for mentions of our target RWM terms. To supplement the manual reading of an article, we searched for keywords through text processing of the article and supplement full text, using the vocabulary assembled in the pilot.

### Data Synthesis and Analysis

The percentages of papers within each epoch using recommended RWM were calculated, along with 95% confidence intervals, and graphed over time. To assess the impact of any article or study characteristics on rates, we conducted a mixed-effects meta-regression using restricted maximum-likelihood (ReML) using *PyMARE*, in Python 3.7 using Jupyter Notebook. [41] Three meta-regressions, one for each phase of research, were done with time as the independent variable. Since the epoch lengths were unequal, we used the midpoint of each epoch in the analysis. To make the intercept meaningful, we subtracted the epoch mid-point (2015) from the epoch year values.

## Sensitivity Analysis

We performed sensitivity analyses for the current study by loosening our categories for phases 1 and 2, to assess the impact of changed definitions. For phase 1 missingness and phase 2 ARWM, we further calculated estimated percentages and their confidence intervals of methods used, using maximal definitions of “assessing missing data” (i.e., giving credit to articles that simply mentioned “missingness” or missing data), and phase 2 ARWM (i.e., adding methods that were not listed by the FDA but still not typical analytical methods; see Appendix Table 2), which defined Combined Missingness and Combined ARWM categories, respectively.

For phase 3, we found no other synonyms for the concept of sensitivity analysis, the third phase of research and thus had no further calculations to perform.

## Results

### Study Selection Flow

The study selection flow is summarized using a PRISMA 2020 flow diagram (Figure 1).[42] The literature search retrieved 5885 citations (for retrieved articles’ distribution, see Appendix Figure 1) We generated a random list of article identifiers for full-text review. We screened the full text of 392 articles total to achieve our target of 35 articles for each epoch. This process resulted in 175 total articles for analysis (for included paper list see Appendix Table 5). Reasons for, and numbers of, exclusions are given in Figure 1.

**Figure 1.**
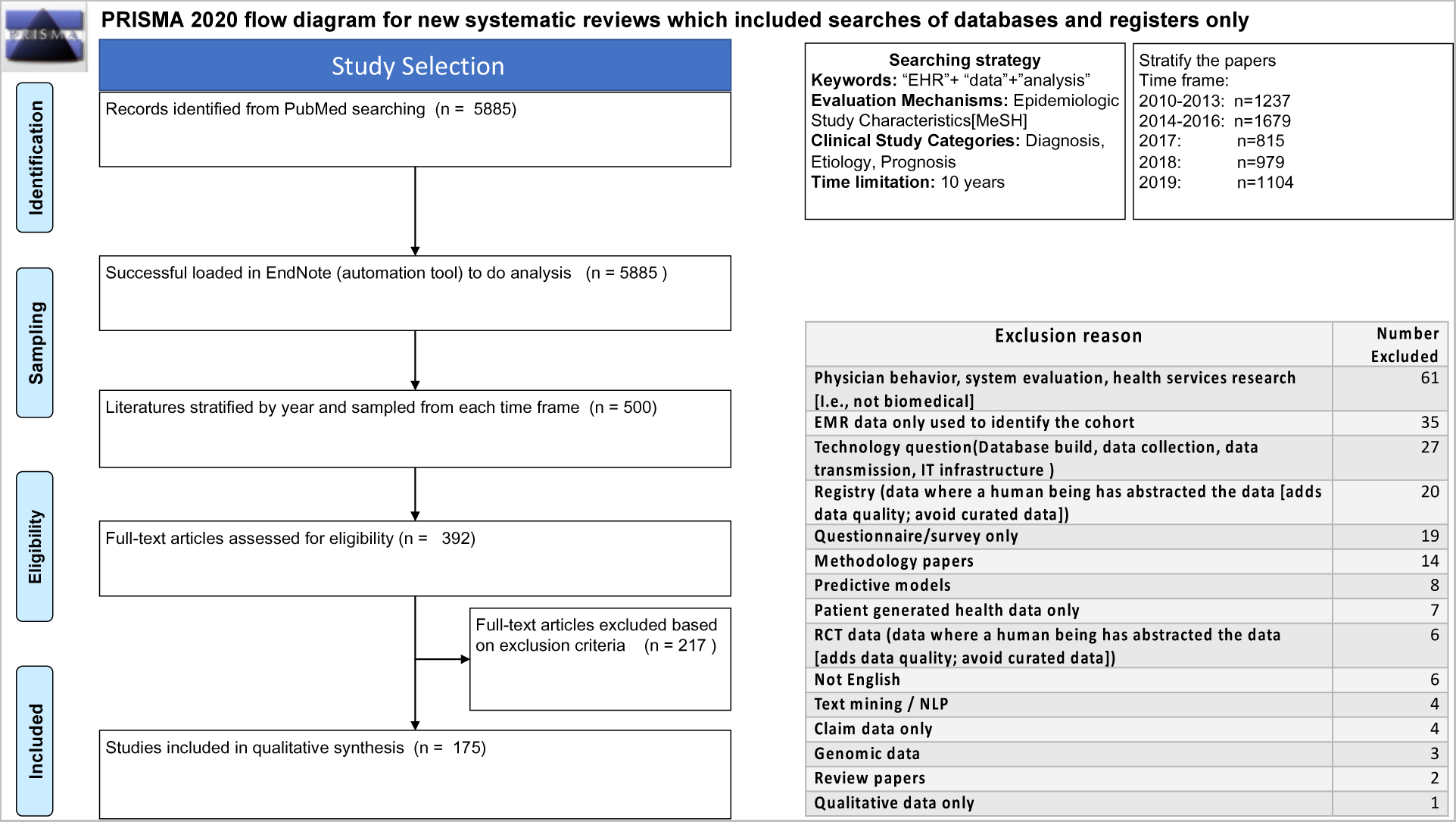
PRISMA Flow Diagram. The characteristics of the 175 included papers (for distribution, see Appendix Figure 2) are provided in Table 2. Of the included papers, 101 (57.7%) were conducted in the North America countries, 37 (21.%) in European countries, and 32 (18.%) in Asia (detailed country distribution see Appendix Table 6). Regarding intention, 142 of the papers aimed at association, and 33, at assessing interventions, with the proportion comparable in each epoch. In terms of design, we characterized 120 (68.6%) as retrospective cohort studies, 14 (8.0%) as retrospective cross-sectional studies, 14 (8.0%) as retrospective chart reviews, and 12 (6.9%) as prospective cohort studies. While a wide variety of statistical tools was reported, 28 (16.0%) reports did not provide such details (detailed statistics tools used see Appendix Table 7). Whereas 4 (2.%) out of 175 papers reported that the research was conducted followed the STROBE[39] check list, no other papers reported using any reporting checklist.

**Table 2.**
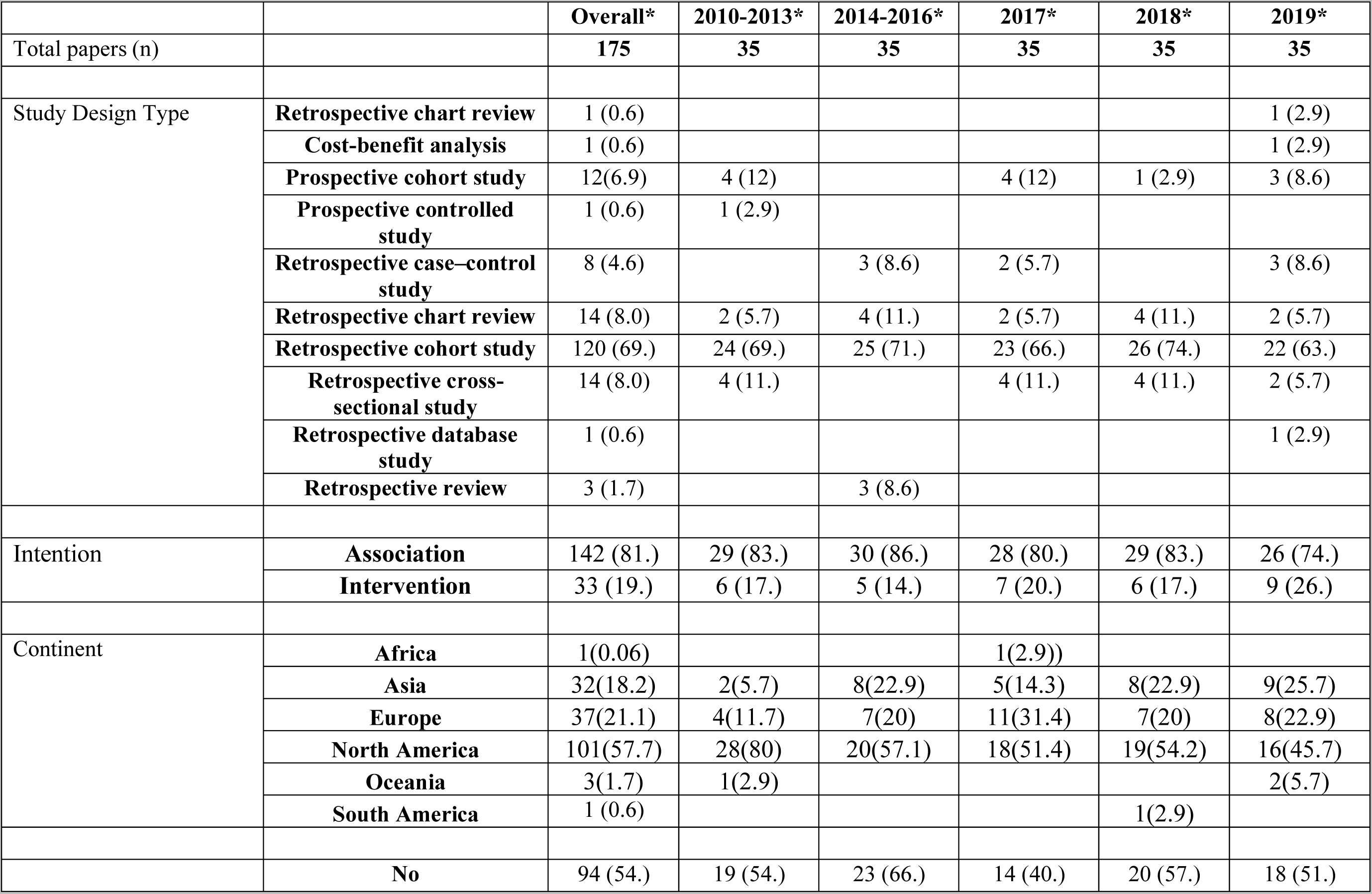
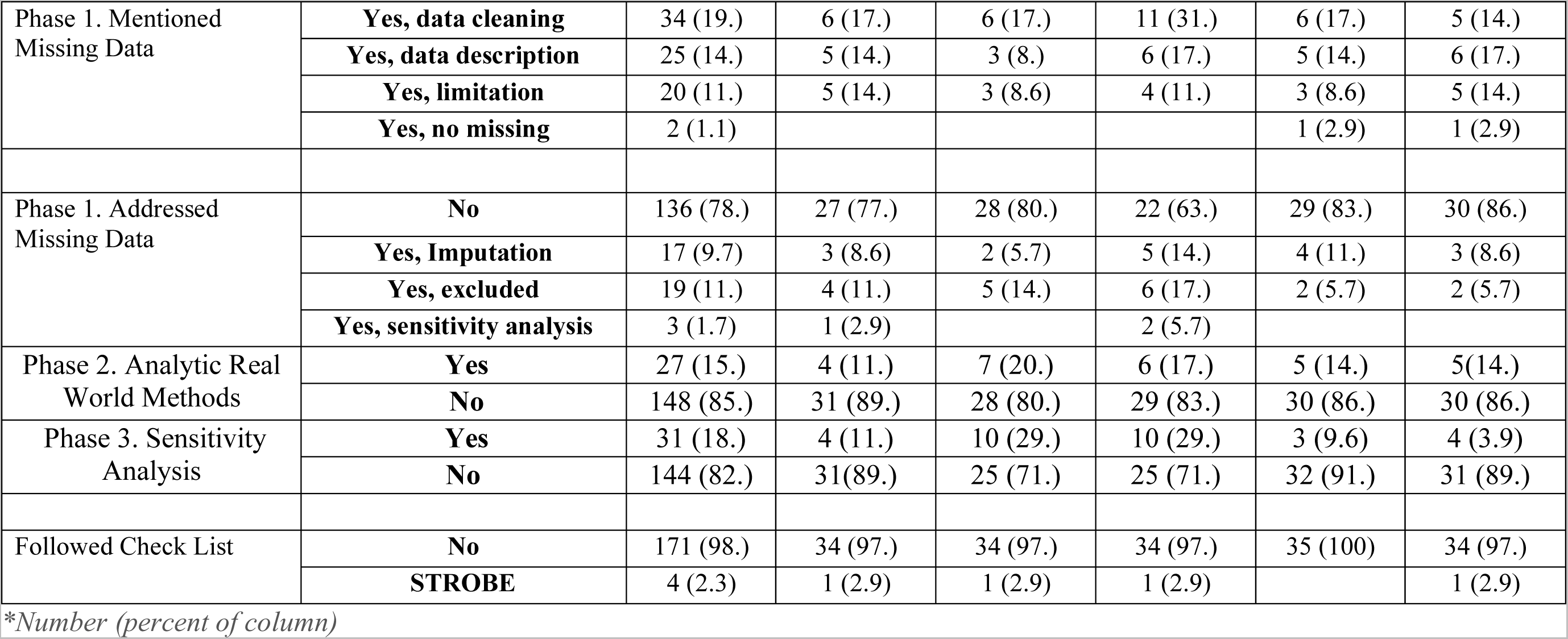
Included Paper Characteristics, by Epoch

### Analysis

Only 1 supplement added the mention of assessed missing data beyond what was found in the main articles. This paper was included in our figures and tables. Thus, for phase 1, the total percentage of articles that assessed missing data was 22%.

Regarding phase 3, the number of papers that reported sensitivity analysis in 2019 is 4 (11%), with a confidence-interval upper bound of 22.%. The range over the 10 years was 12.% to 23.%.

Regarding changes over the years, the raw percentage of recommended methods in each phase, assessing missing data, ARWM, and sensitivity analysis, varied between 10% and 40%. (Figure 2 and Appendix Table 8,) While the proportions seem to rise by 2017, they return to low values in 2018 and 2019.

**Figure 2.**
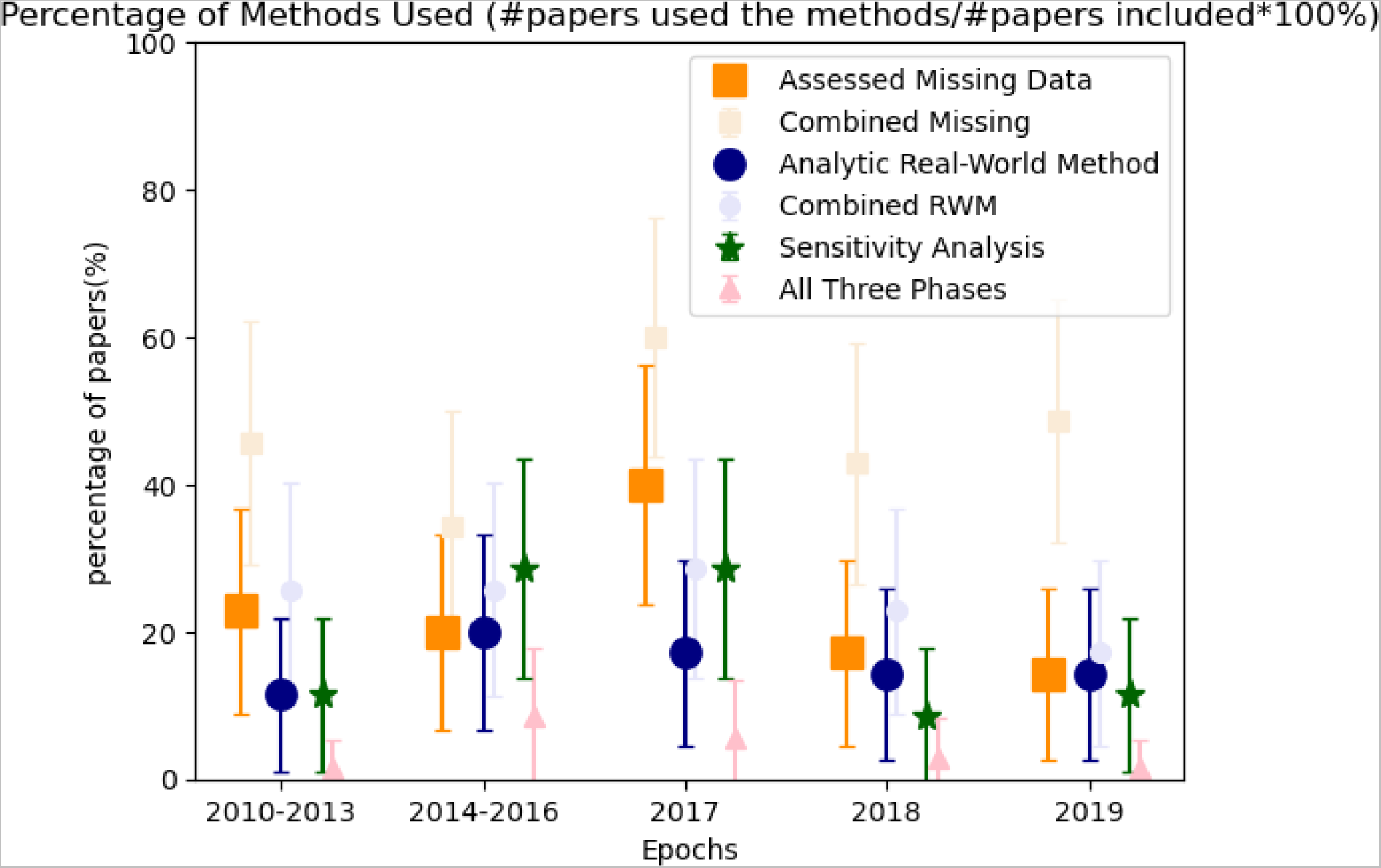
Estimated Percentage of Methods Used in RWD Research, by Epoch. Regarding phase 2, the number of papers that used ARWM in 2019 is 5 (14.%), (with a confidence-interval upper bound of 26.%). Regarding specific traditional methods, 96 (55.%) papers used linear or logistic regressions, 41 papers used survival analysis, with 26 (63. %) not adjusting for baseline or time-varying confounders. [43]

Only 6 papers (3.4%), with a confidence-interval upper bounds of less than 6%, overall addressed recommended RWM in all 3 phases.

### Meta-regressions

The meta-regressions modeled the percentage of method used in each of the three phases as a function of an epoch. The coefficients for the meta-regressions are shown in Table 3. As an example, the coefficient for assessed missing data is −1%, meaning an average drop of 1 percent per epoch after the baseline intercept value of 22% (in 2015) assessing missingness. However, for all other methodology coefficients, the coefficients were close to and statistically equivalent to 0. The percentage of studies using ARWM or using sensitivity analysis did not increase over time.

**Table 3.** Meta-regression for Three Phases

| Variable | Coefficient | SE | z-score | P value | CI Lower Bound | CI Upper Bound |
| --- | --- | --- | --- | --- | --- | --- |
| **Assessed Missing Data over epochs** |  |  |  |  |  |  |
| **Intercept** | 22% | 6.8% | 3.3 | 0.001 | 8.8% | 36% |
| **Epoch** | -1% | 2.1% | -0.34 | 0.73 | -4.7% | 3.3% |
| **Combined Missingness (Sensitivity analysis of Assessed Missing) over epochs** |  |  |  |  |  |  |
| **Intercept** | 45% | 8.1% | 5.5 | 2.9e-08 | 29% | 61% |
| **Epoch** | 0.9% | 2.6% | 0.34 | 0.73 | -4.1% | 5.9% |
| **ARWM used over epochs** |  |  |  |  |  |  |
| **Intercept** | 15% | 5.7% | 2.6 | 0.01 | 3.5% | 26% |
| **Epoch** | 0.3% | 1.7% | 0.18 | 0.86 | -3.1% | 3.7% |
| **Combined ARWM use over epochs (Sensitivity analysis of ARWM use)** |  |  |  |  |  |  |
| **Intercept** | 25% | 7.2% | 3.5 | 5.4e-04 | 11% | 39% |
| **Epoch** | -0.8% | 2.2% | -0.38 | 0.70 | -5.1% | 3.4% |
| **Sensitivity Analysis used over epochs** |  |  |  |  |  |  |
| **Intercept** | 15% | 5.7% | 2.6 | 8.6e-03 | 3.8% | 26% |
| **Epoch** | 0.0% | 1.6% | -0.19 | 0.85 | -3.5% | 2.9% |

*\*Percentage(Factor) = Intercept + Coefficient × (Epoch center - 2015), where Factor = Assessed (Combined) Missing Data, (Combined) Real-World Method, or Performed Sensitivity Analysis* *CI=Confidence interval, ARWM = Analytic Real-World Methods.*

### Sensitivity Analysis

In the case that our categories were too restrictive, we “gave credit” to articles using broader criteria for missingness and RWM definitions. Figure 2 shows the estimated percentages and confidence intervals using these broader criteria (“Combined Missingness” and “Combined ARWM”). The estimates for missingness go as high as 60%, and for ARWM, as high as 29%. As in the meta-regression, there was statistically no improvement over time. Because we found no other synonyms for the concept of sensitivity analysis, we did not pursue the third phase.

## DISCUSSION

We took a novel random-sampling approach for this scoping review to assess the proportion of articles that reported methods recommended for analyzing RWD to generate RWE, over the last 10 years. We divided the RWD-analysis process into 3 phases. The number of papers reporting recommended RWM in all 3 phases was only 6 (3.4%). The proportion of studies using recommended methods within each phase has not changed substantively between 2010 and 2019, whether looking at assessing or just mentioning missing data (phase 1; between 14 and 40%), reporting ARWM (phase 2; between 20 and 40%), or using sensitivity analysis (phase 3; between 9 and 29%). These low rates occurred despite multiple calls by methodology and funding organizations for just such methods throughout this time period.[2,14–16,32,34] While there seems to have been a peak in 2017, the later 2018 and 2019 rates of use of ARWM are no better than in 2010−2013. We biased our inclusion and tagging criteria to be as generous towards use of RWM as possible, so these percentages are probably upper bounds to the true percentages.

The low rates we documented are accompanied by a low documented rate of using methodology checklists. A recent new checklist for reporting RWE—STarT-RWE—includes the requirement to describe how missing data are addressed throughout the analysis and subgroup analyses.[44,45] Of our included papers, 81 (46.%) mentioned a missing-data issue in their data cleaning or limitation section. Of the 81 that did report on missing data, only 39 (48.%) took action beyond mentioning the issue. While statisticians often impute missing data to make maximal use of data that are available, this imputation presumes that missing data are missing at random. However, missed EHR data is generally missing not at random. [21] [46] Methods for missing not at random (MNAR) are more complex and difficult, yet appropriate.[5,47] None of the studies discussed this key concern of EHR data. There are many weaknesses in using EHR data that go beyond the statistical approaches listed in this paper. For instance, improving data quality and accuracy would be paramount, before attempting advanced analytic methods.[48,49] We submit that attention to missing data, ARWM, and sensitivity analysis comprise the minimum of what is needed.

The percentage of papers that used ARWM is low, and remained so for the entire time period, with less than a quarter doing so (although the confidence interval goes as high as 37%). Taking the case of a single, relatively common analysis—survival curves—only 37% used the appropriate method for RWD[26,43]. In addition, the meta regressions suggest that the assessment of missing data, the use of, or the deployment of sensitivity analysis had not changed over 10 years. One could argue that many of the methods listed in Supplementary Table 2 are too recent to expect to appear in our sample. However, “bias,” “adjusting,” “confounding” and “non-adherence” are concerns in epidemiology and evidence-based medicine for decades.[50,51] The low percentage of use of recommended RWM has important implications for the results of the studies: Improper attention to missing data and to selection bias within EHR data means that results from one dataset cannot be applied to other populations in a predictable (and safe) way.[52–54] Improper analytic methods limit generalizability. Lack of sensitivity analysis or other robustness approaches may hide the fragility of the conclusions to contexts outside the RWD dataset.

While we used the FDA meeting[1,2,14–16] as a rich list of modern methods, the sentiment of the need for specific methods to deal with RWD is well established. A standard textbook of Epidemiology calls for bias assessment in “secondary data analysis” (using the definition of RWD). [36] The National Academy of Medicine raises bias issues and results calibration in their 2013 Observational Studies in a Learning Health System workshop summary.[55] The International Society for Pharmacoeconomics and Outcomes Research (ISPOR) and the International Society for Pharmacoepidemiology (ISPE) in 2017 developed recommendations for “Good practices for real-world data studies of treatment and/or comparative effectiveness.” In their discussion, they write:

Threats to the validity of RWD studies of the effects of interventions include unmeasured confounding, measurement error, missing data, model misspecification, selection bias, and fraud. Scholarly journals publish many observational studies, often without requiring the authors to report a thorough exploration of these threats to validity. Unless the authors identify these threats and provide guidance about how they could bias study results, consumers of observational studies must trust the study authors to conduct the study with integrity and report it transparently.[37]

The low percentages we have documented also have important implications for the credibility of EHR-based research. Use of EHRs as a source of data for biomedical research has increased linearly over the past 10 years, rising from 228 biomedical-research articles published based on such data in 2010 to 1104 in 2019 (see Appendix Figure 1). Certainly, such use will only grow, as multi-center efforts, like The National Patient-Centered Clinical Research Network (PCORnet) [56], Observational Health Data Sciences and Informatics (OHDSI)[57] and National COVID Cohort Collaborative (N3C)[58] mature, and as the research community becomes more comfortable with the use of these data. This increase in attention means that it is even more important that proper methods are used for analyzing these data. Furthermore, if the larger corpus of EHR-based research is deemed non-generalizable, biased, and fragile, then the efforts by these organizations may be impugned by the very audience they aim to convince.

One could argue that our study ended too early to see a change a research methodology as a results of guidelines published in 2010s. After all, a 2011 study determined that translating health research takes 17 years.[59] Our results, then, serve as a baseline for RWD research to monitor changes in methodology to satisfy already-published research guidelines.

Because of the complexity of RWD, it is not appropriate to use traditional data processing methods with these large datasets.[60–62] In order to ensure internal and external validity in EHR-based research, researchers must determine whether the data are accurately extracted, adequately adjusted, correctly analyzed and cogently presented.[26] To analyze RWD properly requires multidisciplinary teams comprising clinicians, informaticians, epidemiologists, biostatisticians and data scientists.

The percentage of published studies using recommended RWM has not risen over the past 10 years, despite guidance being issued in multiple years by a variety of sources. Alternative approaches are needed. OHDSI has developed tools to conduct real-world evidence generation.[63] However, at any one institution, a small group of investigators may lack the ability to implement such a sophisticated toolset. Beyond promoting the use of RWM through reporting checklists [64], we suggest a third alternative is to provide analytic decision support, where the analyst receives suggestions from the software in the process of developing an analysis. Examples could include libraries of methods (articles), appropriately tagged; libraries of software code that implements those methods; and metrics to assess successful implementation of the methods. Using the Agency for Healthcare Research and Quality (AHRQ) principle,[65] we suggest that such decision support could do for RWE what EHR-based decision support has done for guideline adherence.[1,66]

## LIMITATIONS

There are several limitations in this research. First, the main data collection was conducted by a single interpreter, which could inject subjectivity into the selection of articles and extraction of significant data. To minimize this potential bias, a uniform approach was adhered to throughout the review process, we had a pilot study that included two reviewers collected data that had a high inter-rater reliability, and a senior reviewer (HL) was consulted in cases of doubt. In addition, the included papers were randomly selected per epoch, providing a representative sample of the literature. Notwithstanding these precautions, it is likely that some important publications were overlooked, or that the sole reviewer impacted the interpretation of the data. Future research may choose to employ several reviewers to boost the validity and applicability of their findings.

Second, we did not include all eligible articles, because our goal was to assess the proportion of such articles using appropriate methods; random sampling is the basis of much biomedical research, our sampling method ensured a representative subset of the literature, allowing for a sound interpretation of the prevalence of recommended RWM usage in the RWD analysis process over the past decade.

Third, we did not include articles from 2020 or later because the COVID-19 pandemic was such a disruption to research practice. Although removing articles published in 2020 or later may limit the scope of our analysis, we made this decision because the COVID-19 pandemic has produced an unprecedented disturbance in research practice. Concerns have also been raised over the quality of research published during the epidemic, as several papers were hurried to publication and lacked sufficient peer review.[67–69] To ensure that our analysis included a more typical sample of studies conducted under regular research conditions, we omitted articles published in 2020 or later. We note, however, that this omission may limit the applicability of our results to more recent studies on RWE approaches. Future research may include analysis of more recent papers and evaluate any modifications to the use of approved RWE procedures considering the pandemic.

Fourth, some EHR-based studies were excluded, such as implementation studies and studies of physician behavior. The analyses of such data are often even more complicated than the more traditional studies of association or medical-intervention effectiveness.[70,71] Certainly, a review of such studies along the lines of the current article would provide a view into how EHR data are used in those contexts as well. We excluded NLP because use of such methods requires informatics expertise. Of 392 articles reviewed, only 4 were rejected because of NLP.

Last but not least, it is possible that not all methodological details were included in the Methods sections which would have led to low observed percentage. However, we reviewed all supplement sections and found only a single additional mention.

## CONCLUSION

The proportion of published clinical research papers based on EHR data using recommended real-world evidence methods is low and has not increased significantly over a 10-year interval. The low percentages we documented may reduce the credibility of EHR-based research, if the larger corpus of EHR-based research is deemed non-generalizable, biased, and fragile. New approaches are needed to promote the use of these methods to improve the validity of studies, to reduce biases, to improve RWE quality, and to better support healthcare decisions. Not doing so risks another 10 years of no improvement in the derivation of evidence from RWD.

## CONTRIBUTORSHIP STATEMENT

HL, CL, and KR designed the study. CL conducted the study including data curation, analysis and interpretation. AA contributed to data curation as a second reviewer. HL reviewed data analysis and interpretation. CL drafted the manuscript that reviewed and revised by HL and KR. implementation.

## Data Availability

https://github.com/ChenyuL/RWE-in-EHR-Data-Analysis

## ACKNOWLEDGEMENTS

We gratefully acknowledge the help of Claire Twose, MLIS, Associate Director for Research Services at Johns Hopkins University, in refining the search strategy and reviewing search implementation. Chenyu Li is supported by University of Pittsburgh School of Medicine Department of Biomedical Informatics Dean’s scholarship.

## Data Availability Statement

The analytic data used for this scoping review was collected during the reviewing process. The retrieved EHR based articles, reviewed article details, and analytic code are available on GitHub at https://github.com/ChenyuL/RWE-in-EHR-Data-Analysis. These resources are provided to facilitate replication and further investigation of the findings presented in this manuscript.

## Appendix and Supplement

### Search Strategy Details (combined)

The following table provides details on the pilot study and the main study search strategies, showing the broader search (all journals) used in the main study.

**Appendix Table 1.**
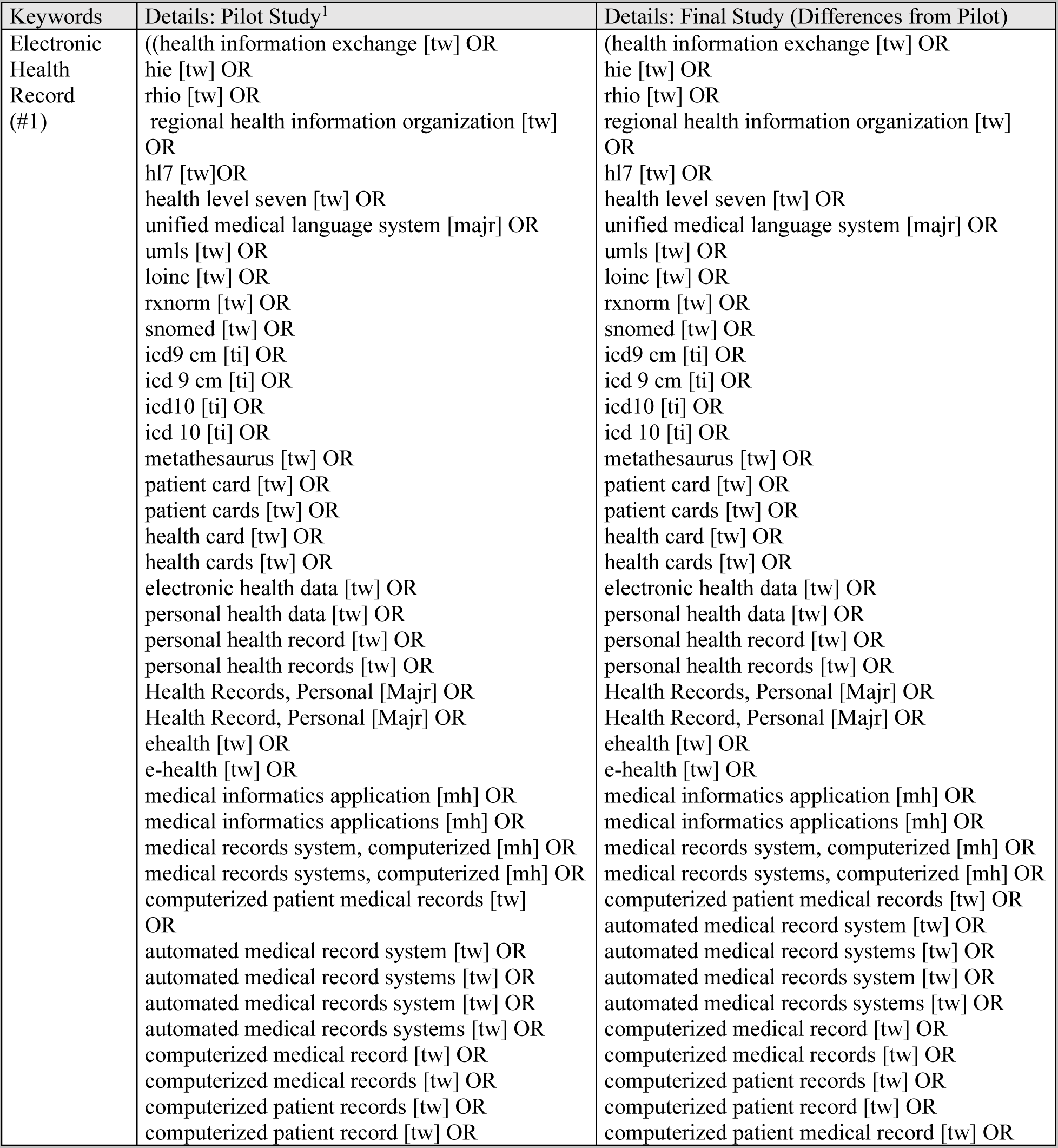
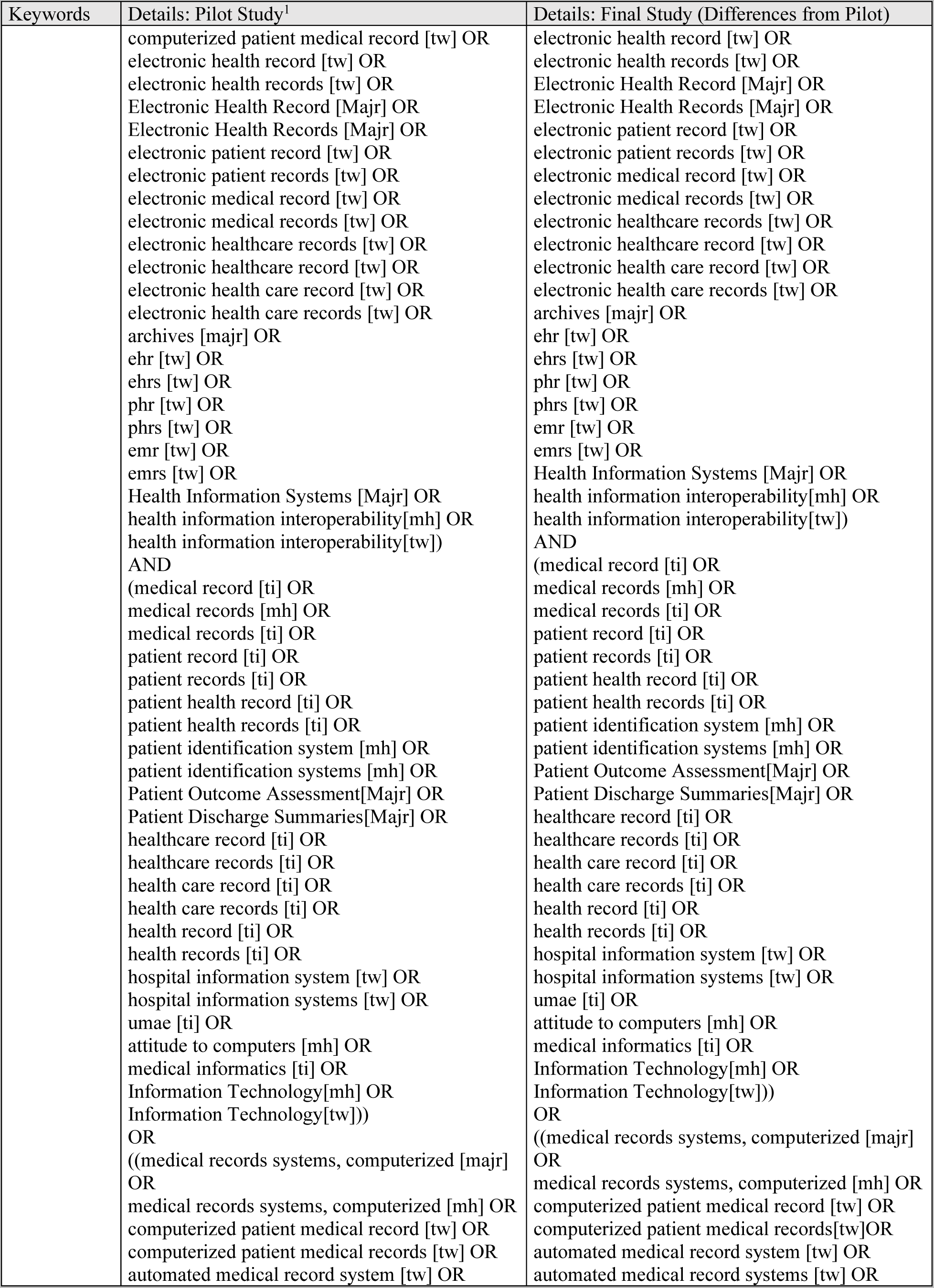
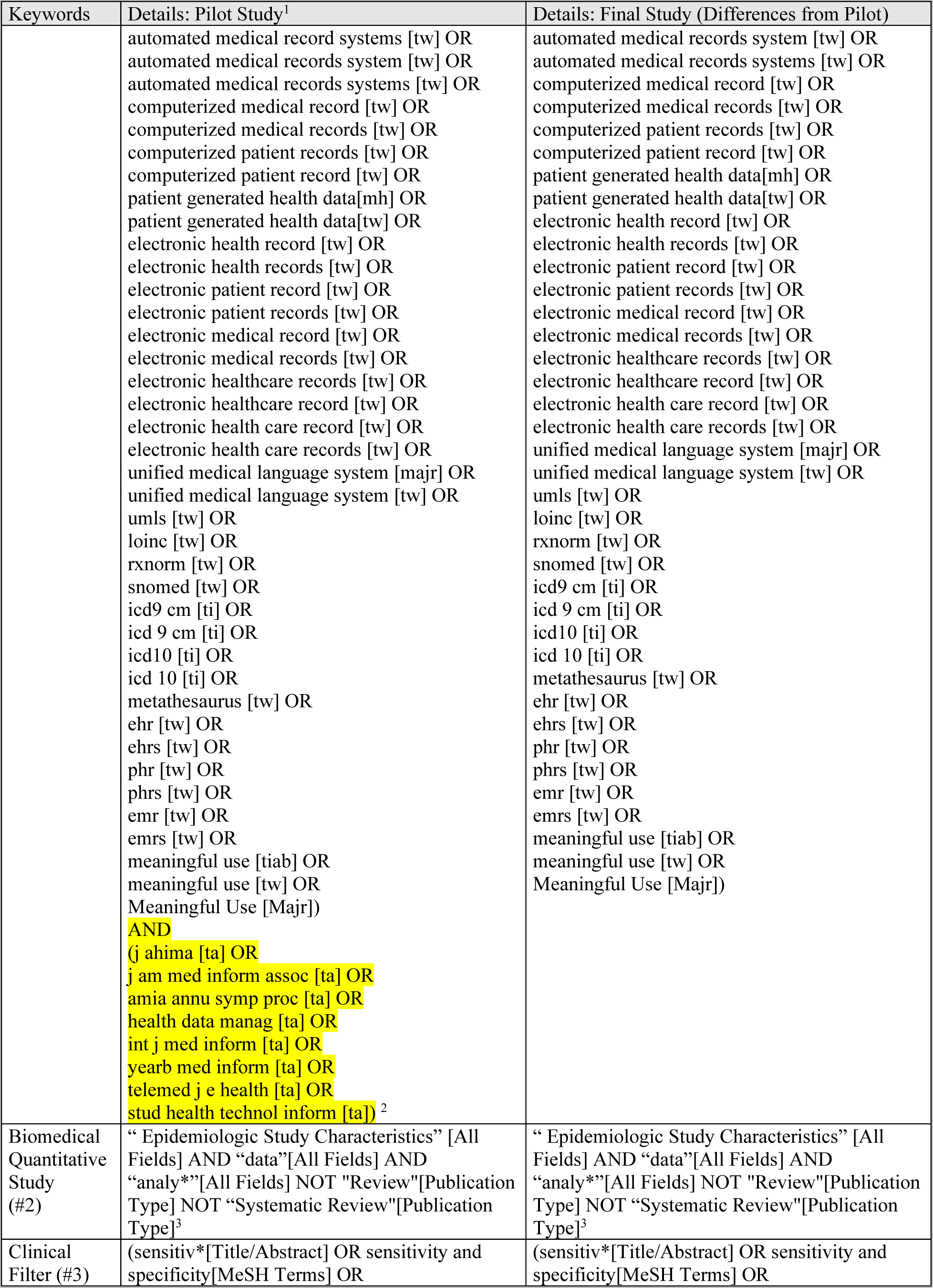
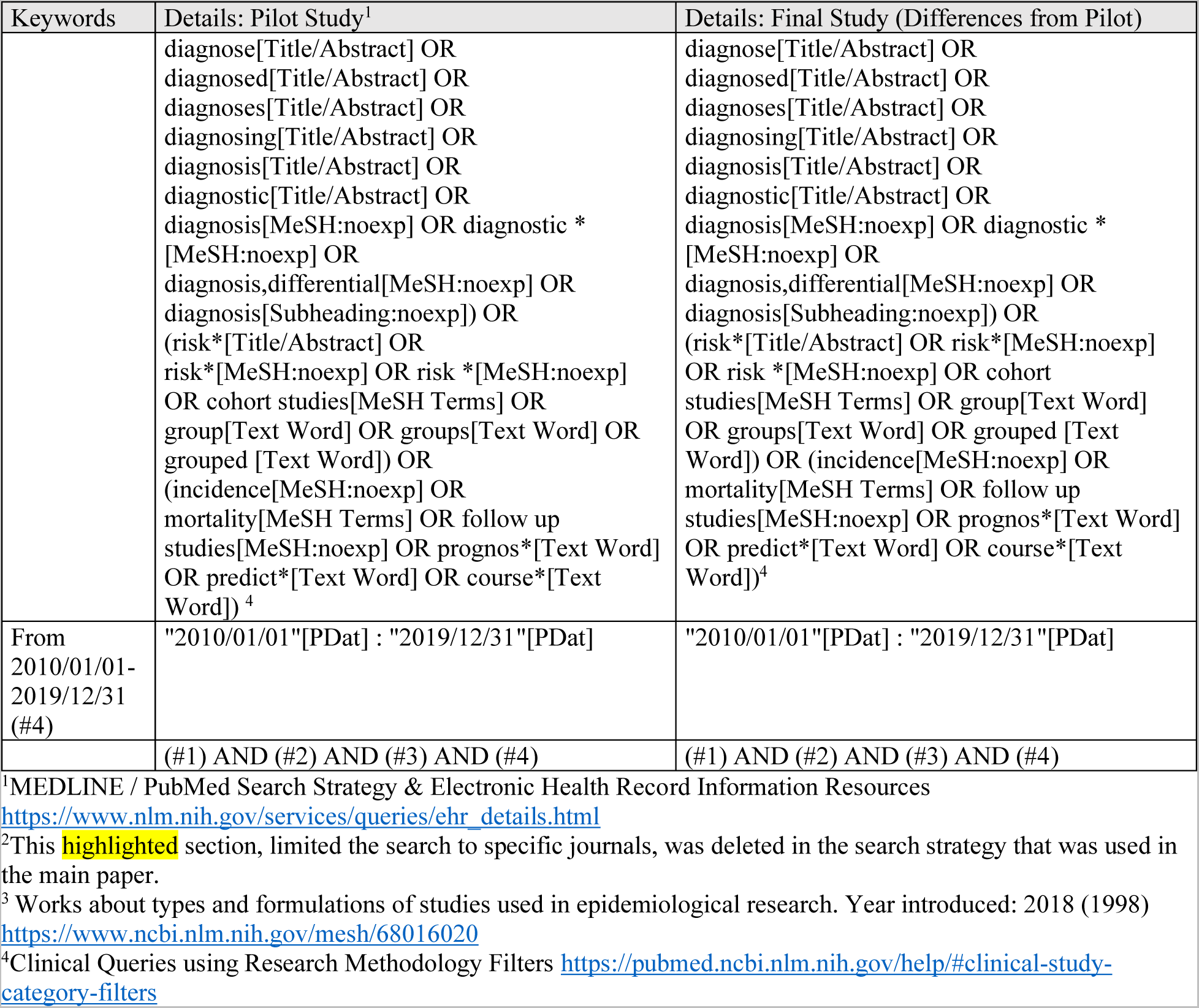
Search Strategies.

| Keywords | Details: Pilot Study<sup>1</sup> | Details: Final Study (Differences from Pilot) |
| --- | --- | --- |
| Electronic Health Record (#1) | ((health information exchange [tw] OR<br>hie [tw] OR<br>rhio [tw] OR<br>regional health information organization [tw]<br>OR<br>hl7 [tw] OR<br>health level seven [tw] OR<br>unified medical language system [majr] OR<br>umls [tw] OR<br>loinc [tw] OR<br>rxnorm [tw] OR<br>snomed [tw] OR<br>icd9 cm [ti] OR<br>icd 9 cm [ti] OR<br>icd10 [ti] OR<br>icd 10 [ti] OR<br>metathesaurus [tw] OR<br>patient card [tw] OR<br>patient cards [tw] OR<br>health card [tw] OR<br>health cards [tw] OR<br>electronic health data [tw] OR<br>personal health data [tw] OR<br>personal health record [tw] OR<br>personal health records [tw] OR<br>Health Records, Personal [Majr] OR<br>Health Record, Personal [Majr] OR<br>ehealth [tw] OR<br>e-health [tw] OR<br>medical informatics application [mh] OR<br>medical informatics applications [mh] OR<br>medical records system, computerized [mh] OR<br>medical records systems, computerized [mh] OR<br>computerized patient medical records [tw]<br>OR<br>automated medical record system [tw] OR<br>automated medical record systems [tw] OR<br>automated medical records system [tw] OR<br>automated medical records systems [tw] OR<br>computerized medical record [tw] OR<br>computerized medical records [tw] OR<br>computerized patient records [tw] OR<br>computerized patient record [tw] OR | (health information exchange [tw] OR<br>hie [tw] OR<br>rhio [tw] OR<br>regional health information organization [tw]<br>OR<br>hl7 [tw] OR<br>health level seven [tw] OR<br>unified medical language system [majr] OR<br>umls [tw] OR<br>loinc [tw] OR<br>rxnorm [tw] OR<br>snomed [tw] OR<br>icd9 cm [ti] OR<br>icd 9 cm [ti] OR<br>icd10 [ti] OR<br>icd 10 [ti] OR<br>metathesaurus [tw] OR<br>patient card [tw] OR<br>patient cards [tw] OR<br>health card [tw] OR<br>health cards [tw] OR<br>electronic health data [tw] OR<br>personal health data [tw] OR<br>personal health record [tw] OR<br>personal health records [tw] OR<br>Health Records, Personal [Majr] OR<br>Health Record, Personal [Majr] OR<br>ehealth [tw] OR<br>e-health [tw] OR<br>medical informatics application [mh] OR<br>medical informatics applications [mh] OR<br>medical records system, computerized [mh] OR<br>medical records systems, computerized [mh] OR<br>computerized patient medical records [tw] OR<br>automated medical record system [tw] OR<br>automated medical record systems [tw] OR<br>automated medical records system [tw] OR<br>automated medical records systems [tw] OR<br>computerized medical record [tw] OR<br>computerized medical records [tw] OR<br>computerized patient records [tw] OR<br>computerized patient record [tw] OR<br>computerized patient medical record [tw] OR |
|  | computerized patient medical record [tw] OR<br>electronic health record [tw] OR<br>electronic health records [tw] OR<br>Electronic Health Record [Majr] OR<br>Electronic Health Records [Majr] OR<br>electronic patient record [tw] OR<br>electronic patient records [tw] OR<br>electronic medical record [tw] OR<br>electronic medical records [tw] OR<br>electronic healthcare records [tw] OR<br>electronic healthcare record [tw] OR<br>electronic health care record [tw] OR<br>electronic health care records [tw] OR<br>archives [majr] OR<br>ehr [tw] OR<br>ehrs [tw] OR<br>phr [tw] OR<br>phrs [tw] OR<br>emr [tw] OR<br>emrs [tw] OR<br>Health Information Systems [Majr] OR<br>health information interoperability[mh] OR<br>health information interoperability[tw])<br>AND<br>(medical record [ti] OR<br>medical records [mh] OR<br>medical records [ti] OR<br>patient record [ti] OR<br>patient records [ti] OR<br>patient health record [ti] OR<br>patient health records [ti] OR<br>patient identification system [mh] OR<br>patient identification systems [mh] OR<br>Patient Outcome Assessment[Majr] OR<br>Patient Discharge Summaries[Majr] OR<br>healthcare record [ti] OR<br>healthcare records [ti] OR<br>health care record [ti] OR<br>health care records [ti] OR<br>health record [ti] OR<br>health records [ti] OR<br>hospital information system [tw] OR<br>hospital information systems [tw] OR<br>umae [ti] OR<br>attitude to computers [mh] OR<br>medical informatics [ti] OR<br>Information Technology[mh] OR<br>Information Technology[tw]))<br>OR<br>((medical records systems, computerized [majr]<br>OR<br>medical records systems, computerized [mh] OR<br>computerized patient medical record [tw] OR<br>computerized patient medical records [tw] OR<br>automated medical record system [tw] OR | electronic health record [tw] OR<br>electronic health records [tw] OR<br>Electronic Health Record [Majr] OR<br>Electronic Health Records [Majr] OR<br>electronic patient record [tw] OR<br>electronic patient records [tw] OR<br>electronic medical record [tw] OR<br>electronic medical records [tw] OR<br>electronic healthcare records [tw] OR<br>electronic healthcare record [tw] OR<br>electronic health care record [tw] OR<br>electronic health care records [tw] OR<br>archives [majr] OR<br>ehr [tw] OR<br>ehrs [tw] OR<br>phr [tw] OR<br>phrs [tw] OR<br>emr [tw] OR<br>emrs [tw] OR<br>Health Information Systems [Majr] OR<br>health information interoperability[mh] OR<br>health information interoperability[tw])<br>AND<br>(medical record [ti] OR<br>medical records [mh] OR<br>medical records [ti] OR<br>patient record [ti] OR<br>patient records [ti] OR<br>patient health record [ti] OR<br>patient health records [ti] OR<br>patient identification system [mh] OR<br>patient identification systems [mh] OR<br>Patient Outcome Assessment[Majr] OR<br>Patient Discharge Summaries[Majr] OR<br>healthcare record [ti] OR<br>healthcare records [ti] OR<br>health care record [ti] OR<br>health care records [ti] OR<br>health record [ti] OR<br>health records [ti] OR<br>hospital information system [tw] OR<br>hospital information systems [tw] OR<br>umae [ti] OR<br>attitude to computers [mh] OR<br>medical informatics [ti] OR<br>Information Technology[mh] OR<br>Information Technology[tw]))<br>OR<br>((medical records systems, computerized [majr]<br>OR<br>medical records systems, computerized [mh] OR<br>computerized patient medical record [tw] OR<br>computerized patient medical records[tw]OR<br>automated medical record system [tw] OR<br>automated medical record systems [tw] OR |
|  | automated medical record systems [tw] OR<br>automated medical records system [tw] OR<br>automated medical records systems [tw] OR<br>computerized medical record [tw] OR<br>computerized medical records [tw] OR<br>computerized patient records [tw] OR<br>computerized patient record [tw] OR<br>patient generated health data[mh] OR<br>patient generated health data[tw] OR<br>electronic health record [tw] OR<br>electronic health records [tw] OR<br>electronic patient record [tw] OR<br>electronic patient records [tw] OR<br>electronic medical record [tw] OR<br>electronic medical records [tw] OR<br>electronic healthcare records [tw] OR<br>electronic healthcare record [tw] OR<br>electronic health care record [tw] OR<br>electronic health care records [tw] OR<br>unified medical language system [majr] OR<br>unified medical language system [tw] OR<br>umls [tw] OR<br>loinc [tw] OR<br>rxnorm [tw] OR<br>snomed [tw] OR<br>icd9 cm [ti] OR<br>icd 9 cm [ti] OR<br>icd10 [ti] OR<br>icd 10 [ti] OR<br>metathesaurus [tw] OR<br>ehr [tw] OR<br>ehrs [tw] OR<br>phr [tw] OR<br>phrs [tw] OR<br>emr [tw] OR<br>emrs [tw] OR<br>meaningful use [tiab] OR<br>meaningful use [tw] OR<br>Meaningful Use [Majr])<br>AND<br>(j ahima [ta] OR<br>j am med inform assoc [ta] OR<br>amia annu symp proc [ta] OR<br>health data manag [ta] OR<br>int j med inform [ta] OR<br>yearb med inform [ta] OR<br>telemed j e health [ta] OR<br>stud health technol inform [ta])<sup>2</sup> | automated medical records system [tw] OR<br>automated medical records systems [tw] OR<br>computerized medical record [tw] OR<br>computerized medical records [tw] OR<br>computerized patient records [tw] OR<br>computerized patient record [tw] OR<br>patient generated health data[mh] OR<br>patient generated health data[tw] OR<br>electronic health record [tw] OR<br>electronic health records [tw] OR<br>electronic patient record [tw] OR<br>electronic patient records [tw] OR<br>electronic medical record [tw] OR<br>electronic medical records [tw] OR<br>electronic healthcare records [tw] OR<br>electronic healthcare record [tw] OR<br>electronic health care record [tw] OR<br>electronic health care records [tw] OR<br>unified medical language system [majr] OR<br>unified medical language system [tw] OR<br>umls [tw] OR<br>loinc [tw] OR<br>rxnorm [tw] OR<br>snomed [tw] OR<br>icd9 cm [ti] OR<br>icd 9 cm [ti] OR<br>icd10 [ti] OR<br>icd 10 [ti] OR<br>metathesaurus [tw] OR<br>ehr [tw] OR<br>ehrs [tw] OR<br>phr [tw] OR<br>phrs [tw] OR<br>emr [tw] OR<br>emrs [tw] OR<br>meaningful use [tiab] OR<br>meaningful use [tw] OR<br>Meaningful Use [Majr]) |
| Biomedical Quantitative Study (#2) | “Epidemiologic Study Characteristics” [All Fields] AND “data” [All Fields] AND “analy*” [All Fields] NOT “Review” [Publication Type] NOT “Systematic Review” [Publication Type]<sup>3</sup> | “Epidemiologic Study Characteristics” [All Fields] AND “data” [All Fields] AND “analy*” [All Fields] NOT “Review” [Publication Type] NOT “Systematic Review” [Publication Type]<sup>3</sup> |
| Clinical Filter (#3) | (sensitiv*[Title/Abstract] OR sensitivity and specificity [MeSH Terms] OR | (sensitiv*[Title/Abstract] OR sensitivity and specificity [MeSH Terms] OR |
|  | diagnose[Title/Abstract] OR<br>diagnosed[Title/Abstract] OR<br>diagnoses[Title/Abstract] OR<br>diagnosing[Title/Abstract] OR<br>diagnosis[Title/Abstract] OR<br>diagnostic[Title/Abstract] OR<br>diagnosis[MeSH:noexp] OR diagnostic *<br>[MeSH:noexp] OR<br>diagnosis,differential[MeSH:noexp] OR<br>diagnosis[Subheading:noexp]) OR<br>(risk*[Title/Abstract] OR<br>risk*[MeSH:noexp] OR risk *[MeSH:noexp]<br>OR cohort studies[MeSH Terms] OR<br>group[Text Word] OR groups[Text Word] OR<br>grouped [Text Word]) OR<br>(incidence[MeSH:noexp] OR<br>mortality[MeSH Terms] OR follow up<br>studies[MeSH:noexp] OR prognos*[Text Word]<br>OR predict*[Text Word] OR course*[Text<br>Word])<sup>4</sup> | diagnose[Title/Abstract] OR<br>diagnosed[Title/Abstract] OR<br>diagnoses[Title/Abstract] OR<br>diagnosing[Title/Abstract] OR<br>diagnosis[Title/Abstract] OR<br>diagnostic[Title/Abstract] OR<br>diagnosis[MeSH:noexp] OR diagnostic *<br>[MeSH:noexp] OR<br>diagnosis,differential[MeSH:noexp] OR<br>diagnosis[Subheading:noexp]) OR<br>(risk*[Title/Abstract] OR risk*[MeSH:noexp]<br>OR risk *[MeSH:noexp] OR cohort<br>studies[MeSH Terms] OR group[Text Word]<br>OR groups[Text Word] OR grouped [Text<br>Word]) OR (incidence[MeSH:noexp] OR<br>mortality[MeSH Terms] OR follow up<br>studies[MeSH:noexp] OR prognos*[Text Word]<br>OR predict*[Text Word] OR course*[Text<br>Word])<sup>4</sup> |
| From<br>2010/01/01-2019/12/31<br>(#4) | "2010/01/01"[PDat] : "2019/12/31"[PDat] | "2010/01/01"[PDat] : "2019/12/31"[PDat] |
|  | (#1) AND (#2) AND (#3) AND (#4) | (#1) AND (#2) AND (#3) AND (#4) |

<sup>1</sup>MEDLINE / PubMed Search Strategy & Electronic Health Record Information Resources
https://www.nlm.nih.gov/services/queries/ehr_details.html
<sup>2</sup>This highlighted section, limited the search to specific journals, was deleted in the search strategy that was used in the main paper.
<sup>3</sup> Works about types and formulations of studies used in epidemiological research. Year introduced: 2018 (1998)
https://www.ncbi.nlm.nih.gov/mesh/68016020
<sup>4</sup>Clinical Queries using Research Methodology Filters https://pubmed.ncbi.nlm.nih.gov/help/#clinical-study-category-filters

During the pilot review, we identified several issues with that search strategy, for instance it was limited to 8 informatics journals, while its terms for “Biomedical Quantitative Study” were too broad1^2^. In the main study, we removed the journal restrictions on NLM-supplied Electronic Health Records search strategy, details see Appendix Table 1. “Biomedical Research” was operationalized as the PubMed Publication Type, “Study Characteristics.” To limit the result to quantitative biomedical studies with data analytic methods, we added keywords “data”[All Fields] AND “analy*”[All Fields] to the search. We used PubMed clinical filters to focus on diagnosis, etiology, prognosis studies, the broad definition for searching diagnosis, etiology, and prognosis has sensitivity of 90%, 93%, and 90%, respectively.^3^

The explanation provided in Appendix 2 of our manuscript refers to the process we used to categorize Real-World Methods (RWM) and Traditional Methods for dealing with missing data in EHR-based biomedical research.

Specifically, when we reviewed the articles, we looked for any mention of a method being used for RWD analysis. If a method was specifically mentioned in the context of RWD analysis, we considered it to be an RWM and placed it on the left side of the table in Appendix 2.

However, if a method was not explicitly mentioned in the context of RWD analysis, we considered it to be a Traditional Method and placed it on the right side of the table.

For example, in the case of logistic regression, while it is often used in RWD analysis, it can also be used in traditional data analysis. Therefore, if it was not explicitly mentioned in the context of RWD analysis, we considered it to be a Traditional Method and placed it on the right side of the table.

It is important to note that our categorization of methods as RWM or Traditional Methods was based on our interpretation of the articles and the information provided by the authors. It is possible that other researchers may categorize these methods differently, based on their own interpretation of the literature or other criteria.

Overall, the purpose of Appendix 2 was to provide a comprehensive list of methods used in EHR-based biomedical research, categorized according to whether they were RWM or Traditional Methods. This information can be used by researchers to select appropriate methods for their own studies and to compare the methods used in different studies.

**Appendix Table 2.** Real World Methods List.

| Phase | Real World Methods (RWM) | Traditional Methods |
| --- | --- | --- |
| Missingness | Missing Cause<br>Missing Data<br>Missingness<br>Multiple Imputation |  |
| Analytic Real World Method | Active Comparator<br>Adjusting<br>Bayesian Twin Regression<br>Bias<br>Causal Inference<br>Confounding<br>DAG/ADMG<br>Depletion of Susceptible<br>Difference in Difference<br>Doubly Robust Methods<br>Empirical Calibration<br>G-Estimation<br>High-dimensional Proxy Adjustment<br>Identification<br>Immortal Time<br>Instrumental Variable<br>Inverse Probability<br>Latent Class Growth Modeling (LCGM)<br>(Manski's) Partial Identification<br>Marginal Structure Models<br>Negative Control<br>Non-adherence<br>Perturbation Variable<br>Propensity Score<br>Pseudo Treatment<br>Regression Discontinuity<br>Reverse Causation<br>Targeted Maximum Likelihood Estimation<br>Trend in Trend<br>Trimming | Adaboost *<br>C-Statistics<br>Chi-Square Automatic Interaction Detector<br>Cox proportional hazard models<br>Decision Curve Analysis *<br>Descriptive statistics<br>Difference-in-difference analysis<br>Discriminant function analysis<br>Empirical Bayes estimates*<br>Gamma Regression<br>Generalized Estimating Equations *<br>Group-based multitrajectory analysis *<br>Growth mixture modelling *<br>Hierarchic cluster analysis *<br>Hierarchical generalized linear model *<br>Hypothesis test<br>Individual growth curve (IGC) analysis*<br>Intention-to-Treat Analysis<br>Interrupted time series analysis<br>Kaplan-Meier method<br>Latent Class Analysis *<br>LASSO regression *<br>Life-table analysis<br>Linear mixed effect model<br>Linear Regression<br>Logistic Regression<br>Long Short Term Memory (LSTM) Network<br>Mixed -effects Regression Model<br>Multilevel logistic regression *<br>Multilevel quantile regression<br>Multiple correspondence analysis<br>Multiple logistic regression<br>Multivariable generalized linear mixed model<br>Multivariable negative binomial regression<br>Multivariate analysis of variance (MANOVA)<br>Multivariate linear regression<br>Multivariate logistic regression<br>One-way analysis of variance (ANOVA)<br>Optimal Classification Trees *<br>Poisson regression *<br>Post-hoc analysis<br>Random Forest *<br>Recursive partitioning (RP) model *<br>Retrospective chart review *<br>Statistic Testing<br>Subgroup Analysis |
| Sensitivity Analysis | Sensitivity Analysis |  |

### Pilot Study Results

We sampled 300 papers in order to gain a stable estimate of our target proportions

**Appendix Table 3.** Number of articles retrieved, by year.

| Year | 2010-2013 | 2014-2016 | 2017 | 2018 | 2019 |
| --- | --- | --- | --- | --- | --- |
| Total papers | 350 | 341 | 157 | 189 | 91 |
| Proportion of total papers | 31.0% | 30.2% | 13.9% | 16.8% | 8.1% |
| Sampled for full-text reading | 100 | 50 | 50 | 50 | 50 |

**Appendix Table 2.**
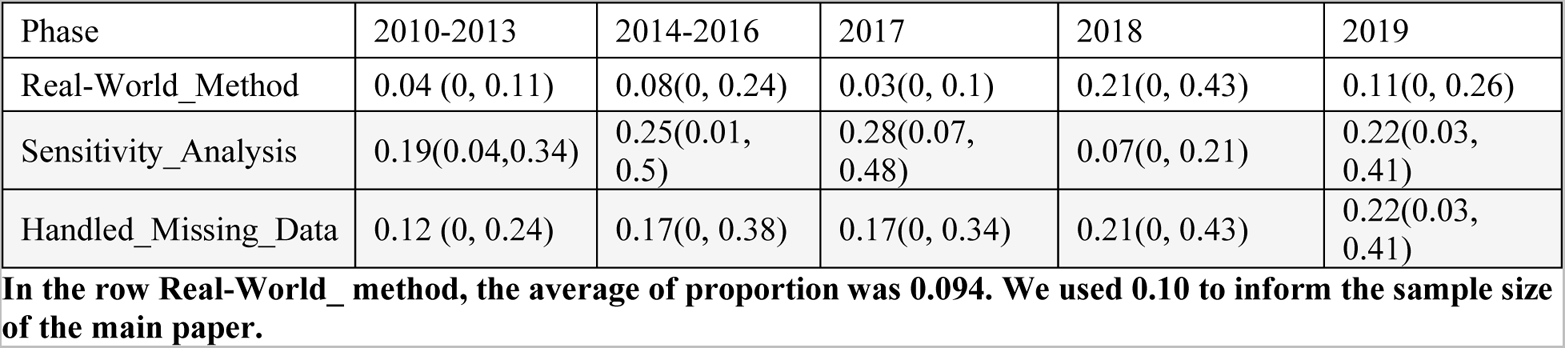
Pilot Study Estimated Proportions and Confidence Interval of Methods used in EHRs Based Research.

| Phase | 2010-2013 | 2014-2016 | 2017 | 2018 | 2019 |
| --- | --- | --- | --- | --- | --- |
| Real-World_Method | 0.04 (0, 0.11) | 0.08(0, 0.24) | 0.03(0, 0.1) | 0.21(0, 0.43) | 0.11(0, 0.26) |
| Sensitivity_Analysis | 0.19(0.04,0.34) | 0.25(0.01, 0.5) | 0.28(0.07, 0.48) | 0.07(0, 0.21) | 0.22(0.03, 0.41) |
| Handled_Missing_Data | 0.12 (0, 0.24) | 0.17(0, 0.38) | 0.17(0, 0.34) | 0.21(0, 0.43) | 0.22(0.03, 0.41) |

In the row Real-World\_ method, the average of proportion was 0.094. We used 0.10 to inform the sample size of the main paper.

### Main Study Results

**Appendix Figure 1:**
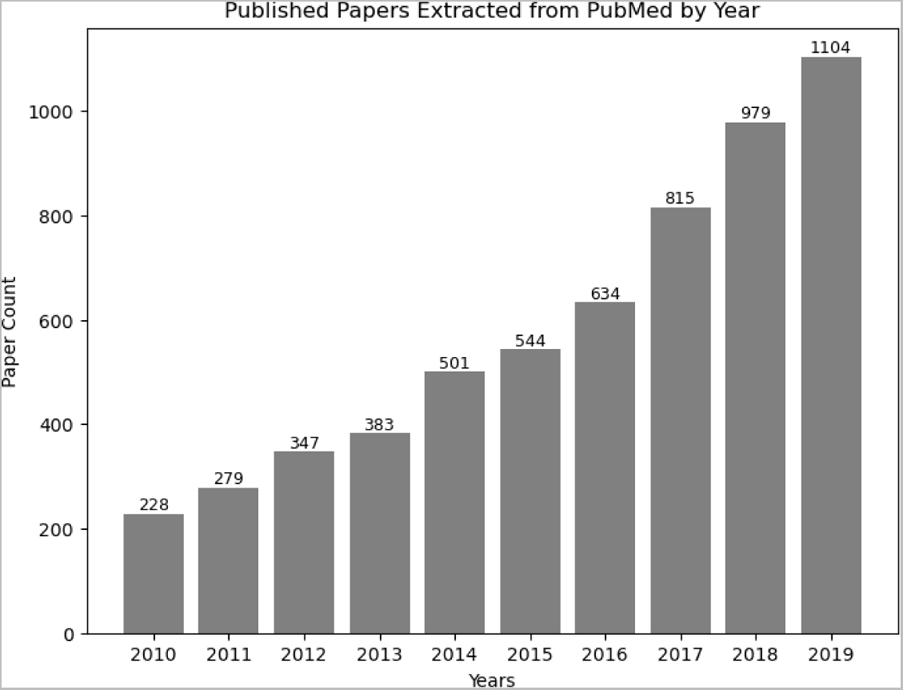
Published Papers Extracted from PubMed, by Year

**Appendix Figure 1.**
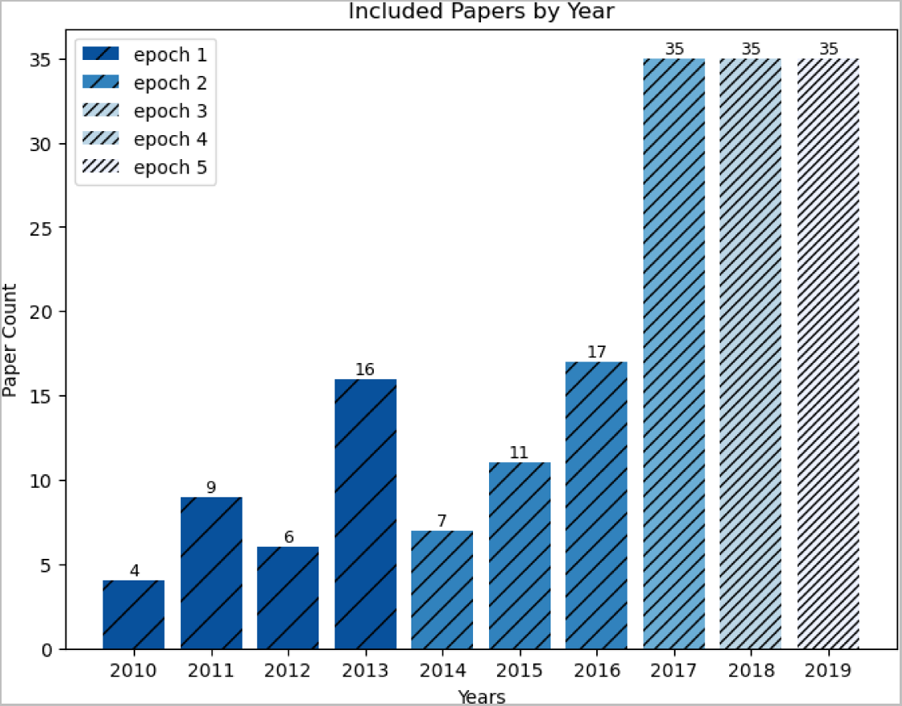
Included Papers, by Year

**Appendix Table 5.**
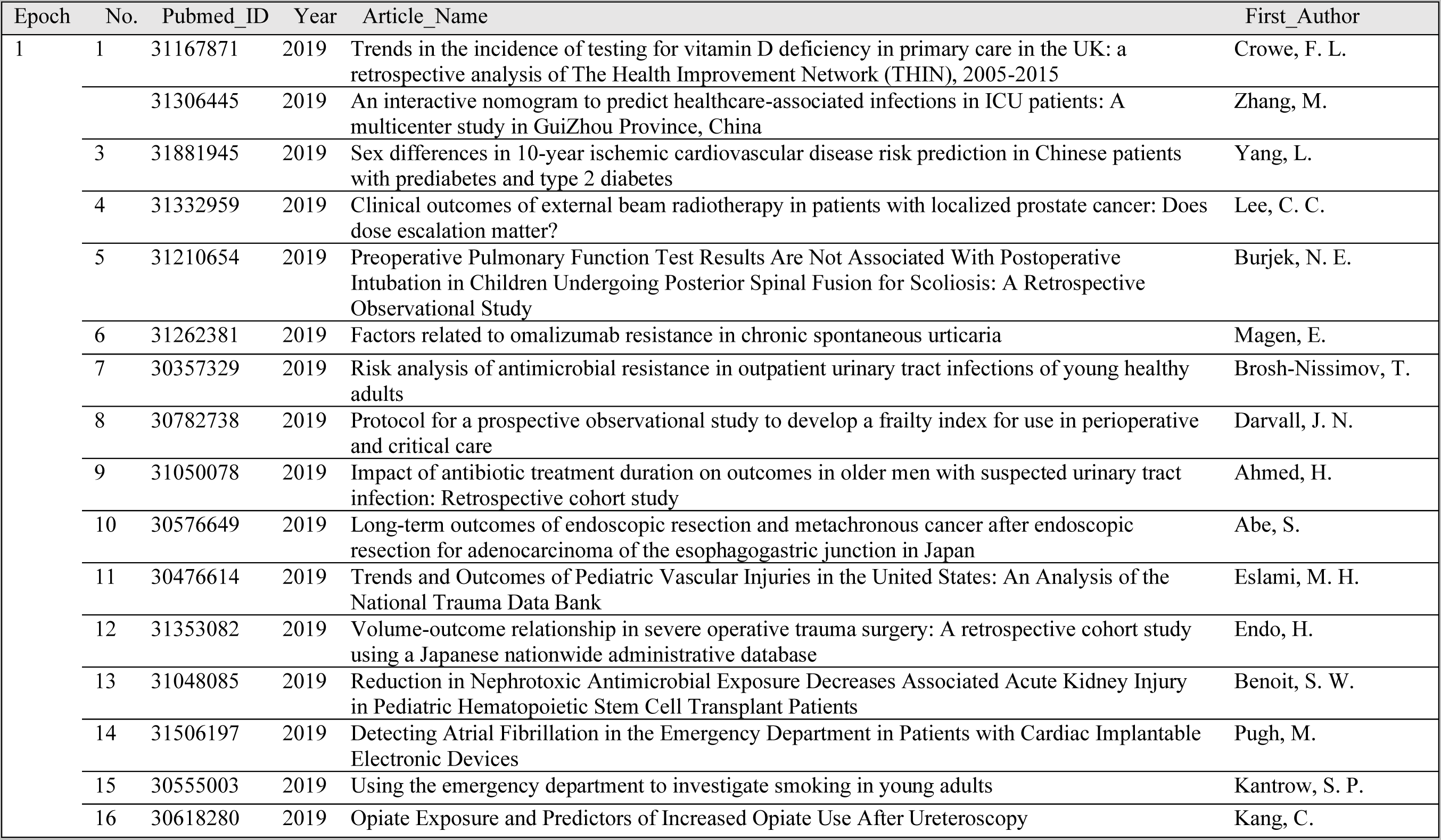
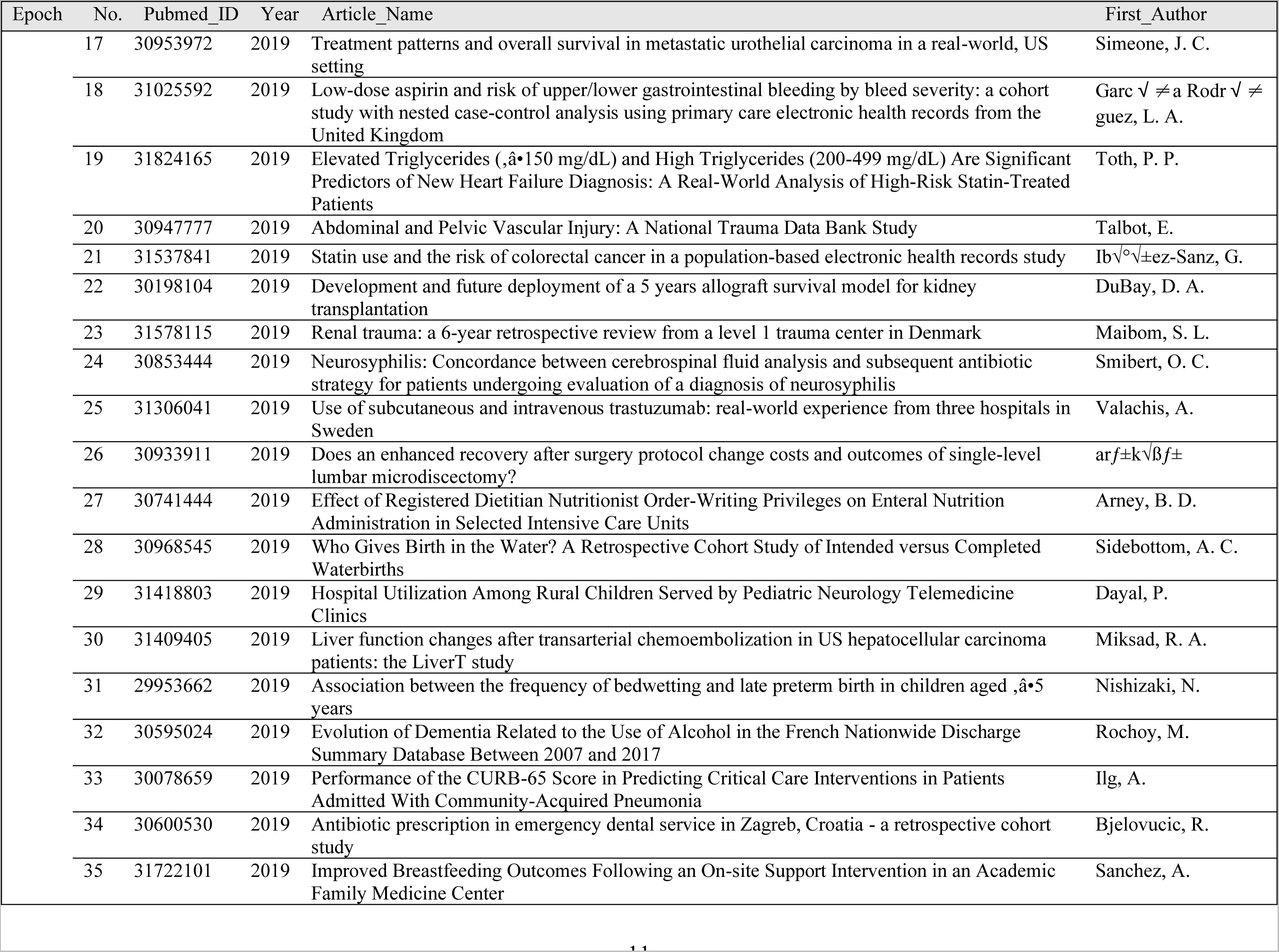
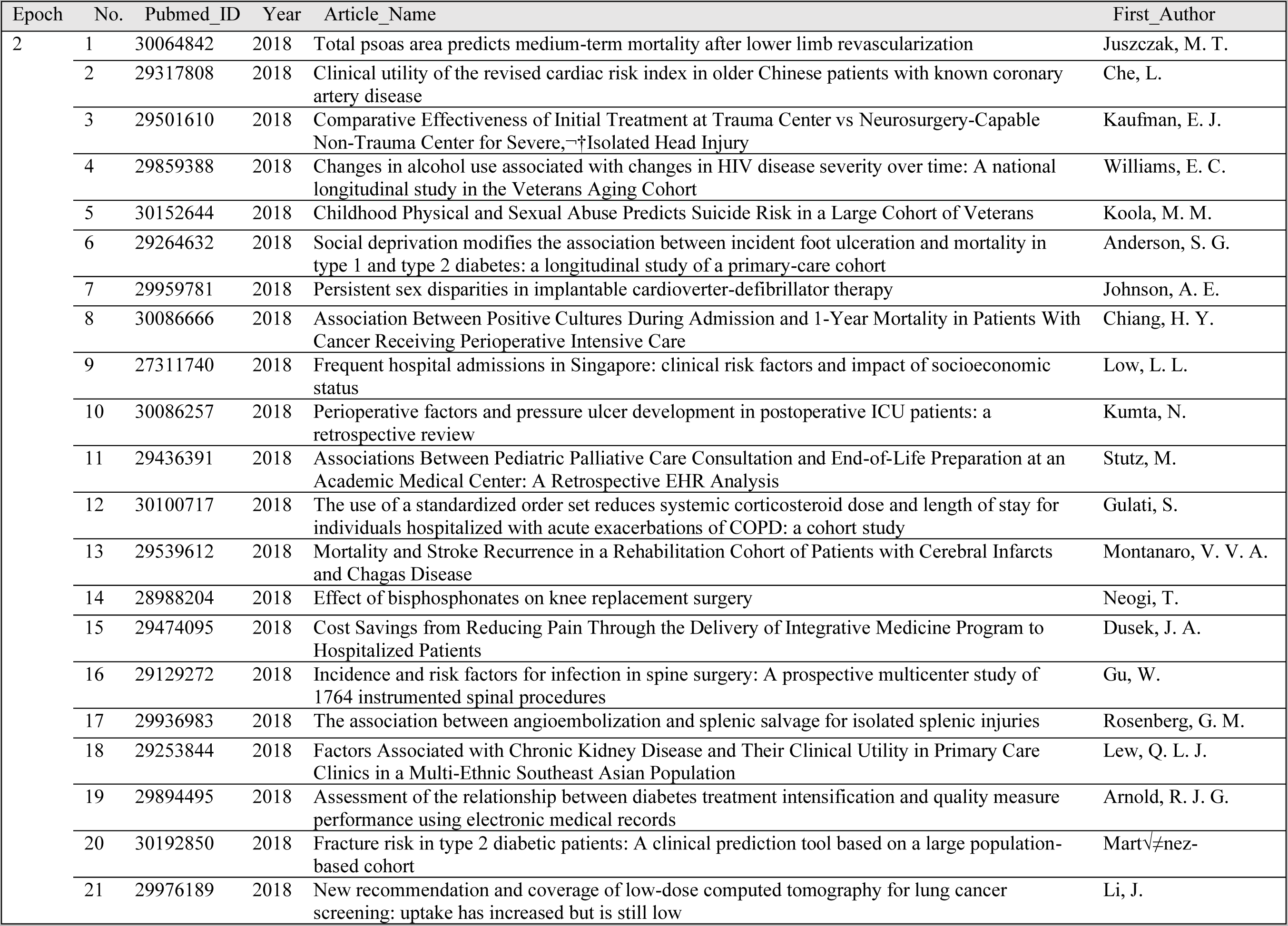
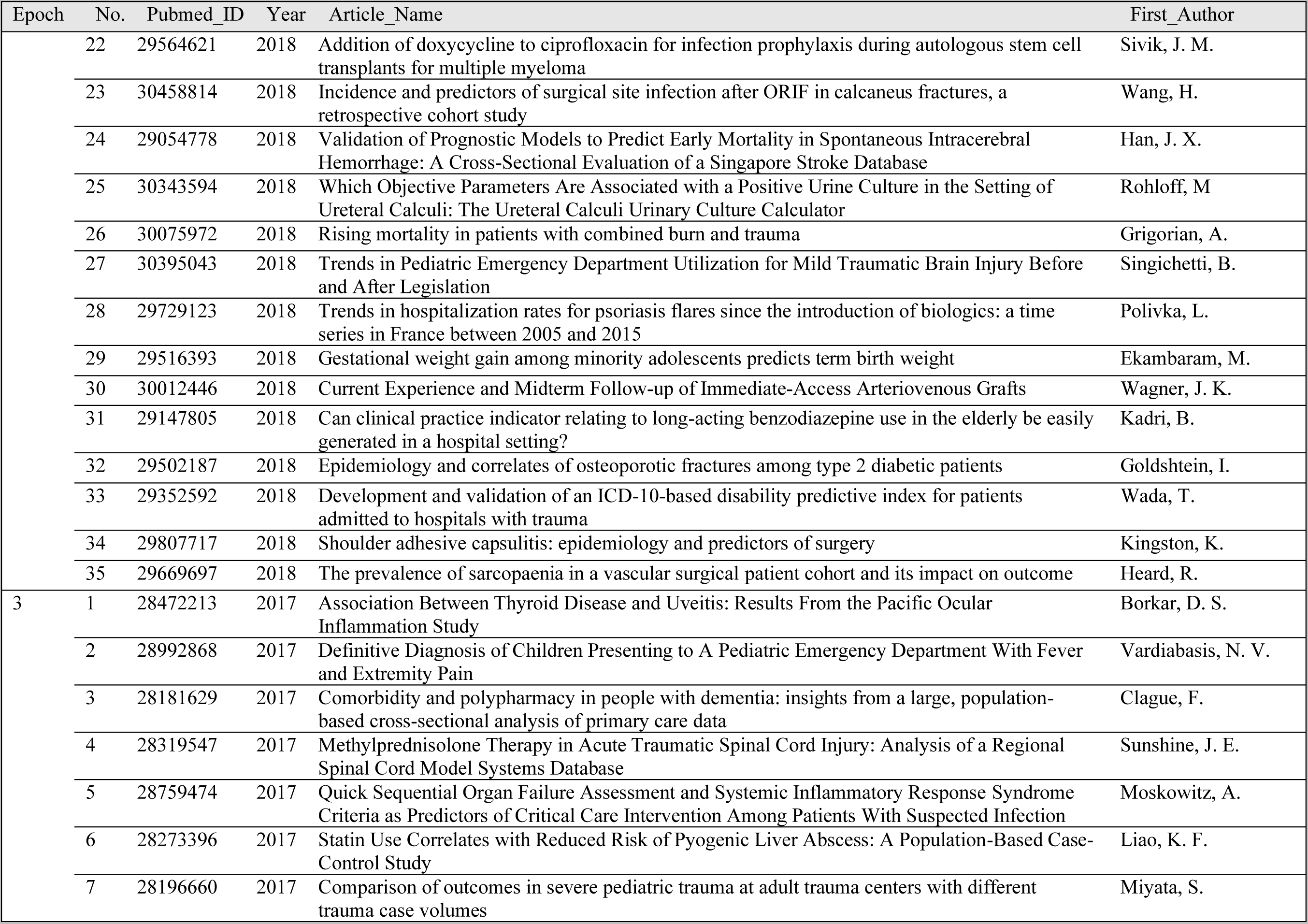
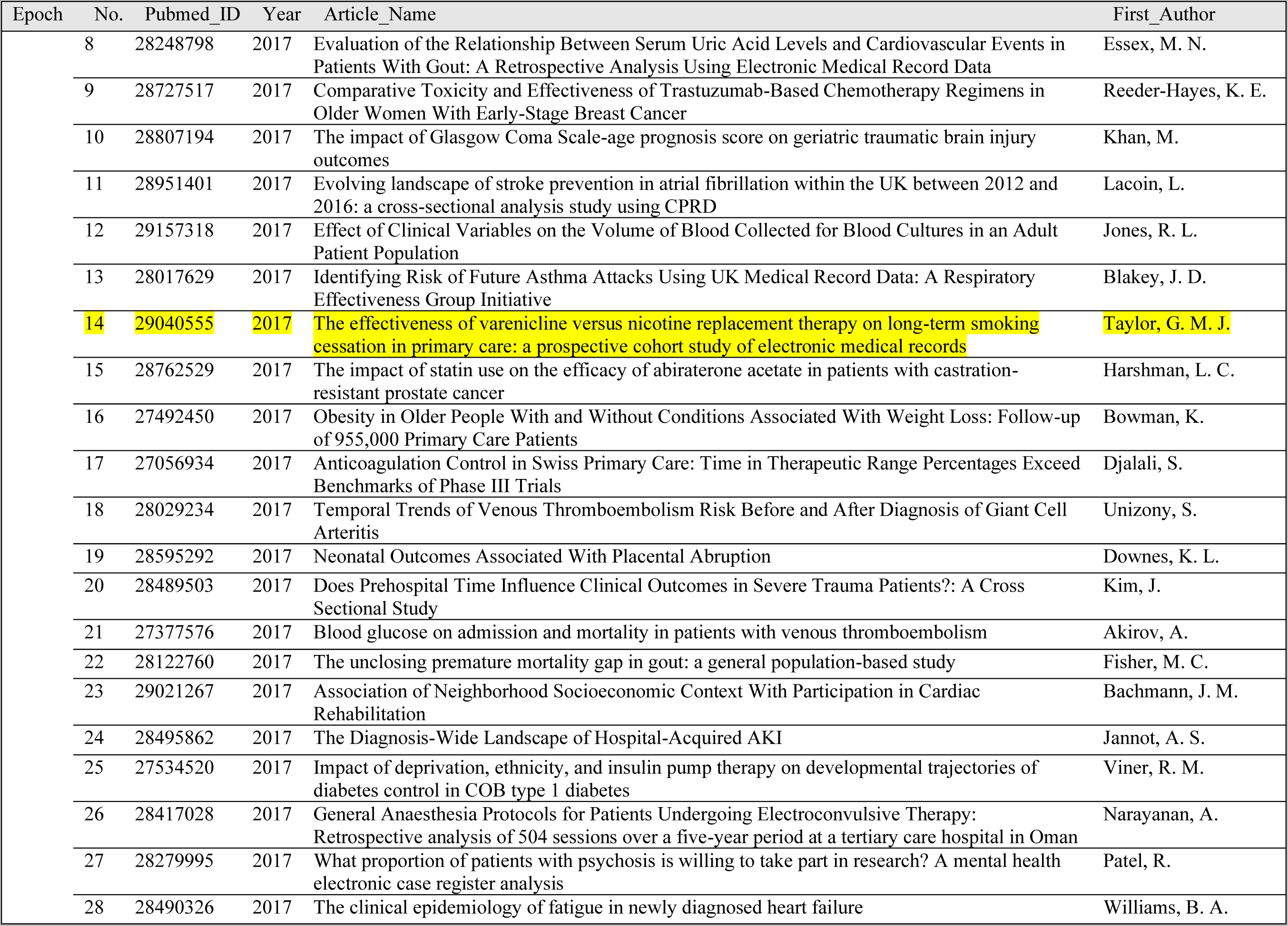
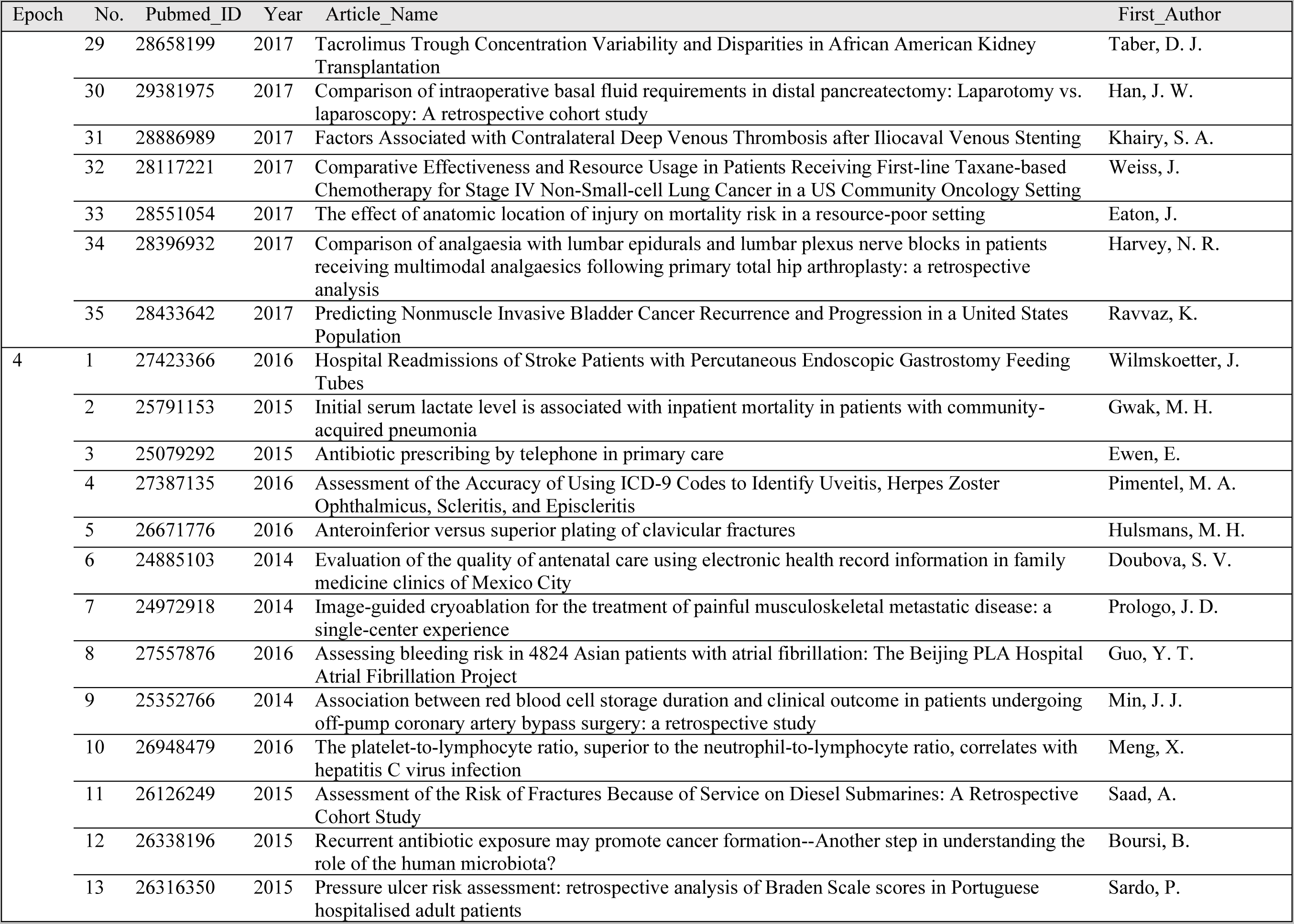
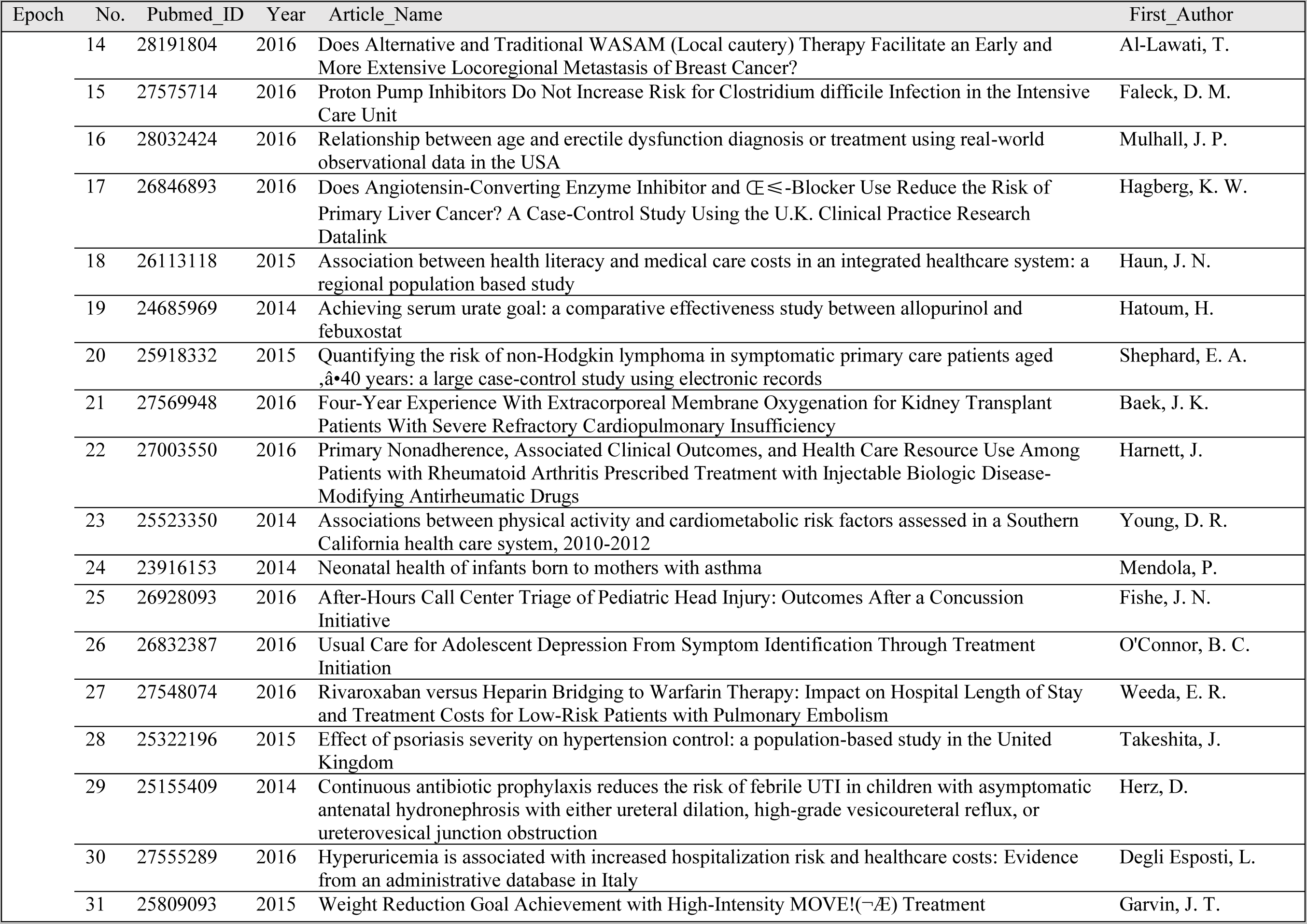
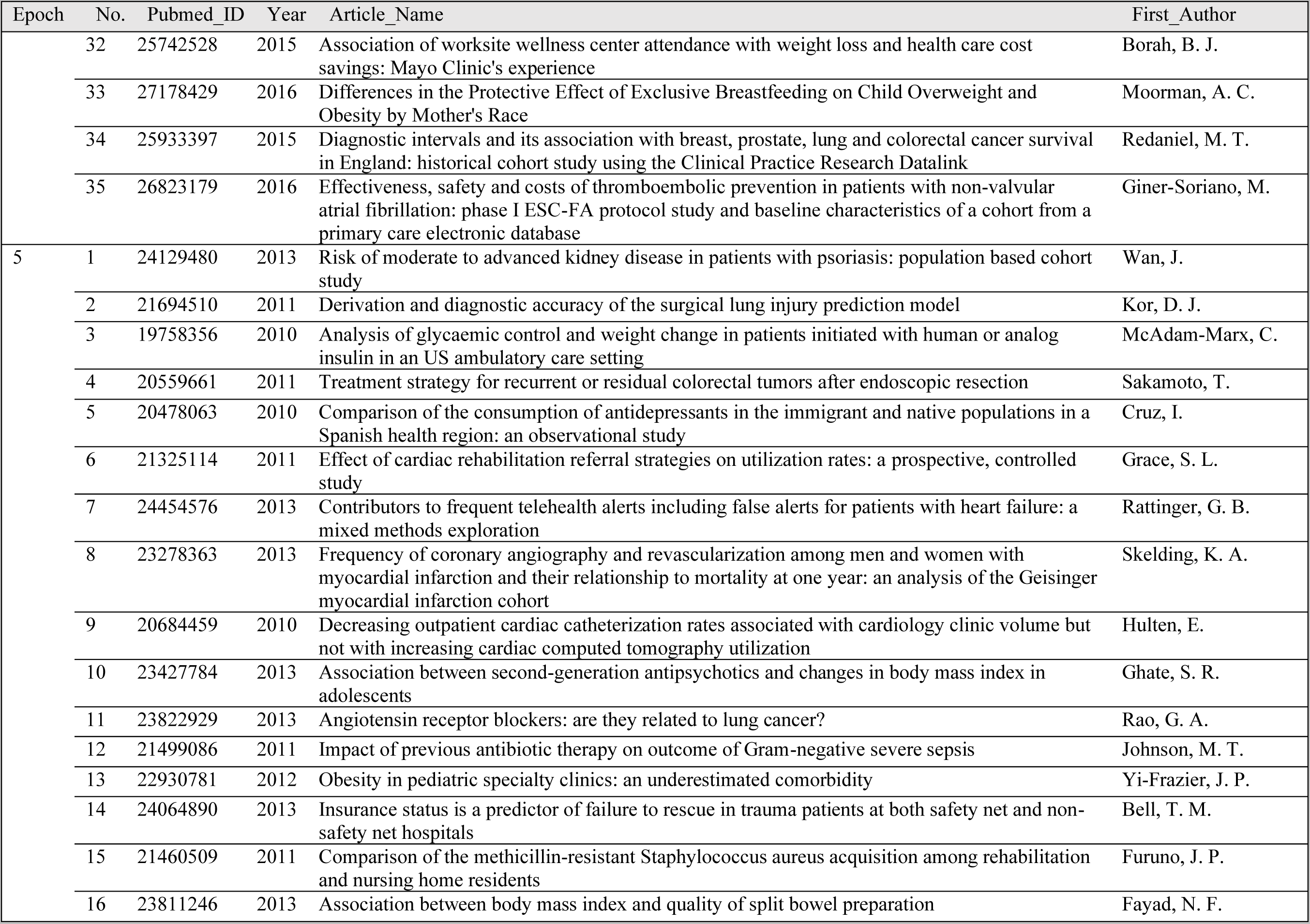
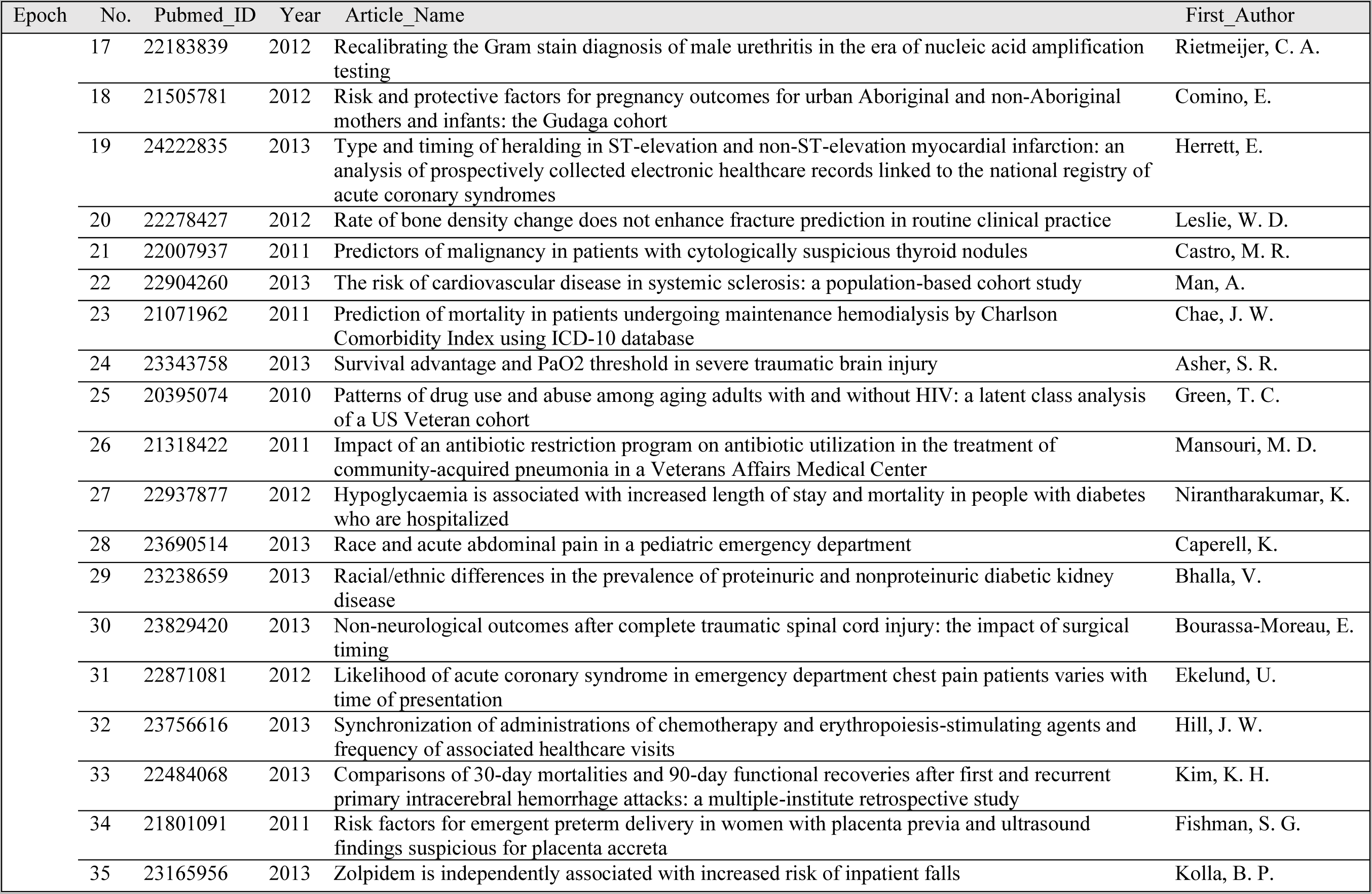
Reviewed Paper List^4^.

**Appendix Table 6.** Main Study Included Paper Countries Details, by Epoch.

Details, by Epoch
| Country/district | Overall* | 2010-2013* | 2014-2016* | 2017* | 2018* | 2019* |
| --- | --- | --- | --- | --- | --- | --- |
|  | 175(100) | 35(100) | 35(100) | 35(100) | 35(100) | 35(100) |
| Australia | 3 (1.7) | 1 (2.9) |  |  |  | 2 (5.7) |
| Brazil | 1 (0.6) |  |  |  | 1 (2.9) |  |
| Canada | 3 (1.7) | 3 (8.6) |  |  |  |  |
| China | 7 (4.0) |  | 2 (5.7) |  | 3 (8.6) | 2 (5.7) |
| Croatia | 1 (0.6) |  |  |  |  | 1 (2.9) |
| Denmark | 1 (0.6) |  |  |  |  | 1 (2.9) |
| France | 5 (2.9) |  |  | 2 (5.7) | 2 (5.7) | 1 (2.9) |
| Israel | 6 (3.4) |  | 2 (5.7) | 1 (2.9) | 1 (2.9) | 2 (5.7) |
| Italy | 1 (0.6) |  | 1 (2.9) |  |  |  |
| Japan | 4 (2.3) |  |  |  | 1(2.9) | 3 (8.6) |
| Korea | 7 (4.0) | 2 (5.7) | 3 (8.6) | 2 (5.7) |  |  |
| Malawi | 1 (0.6) |  |  | 1 (2.9) |  |  |
| Mexico | 1 (0.6) |  | 1 (2.9) |  |  |  |
| Netherlands | 1 (0.6) |  | 1 (2.9) |  |  |  |
| Oman | 2 (1.1) |  | 1 (2.9) | 1 (2.9) |  |  |
| Portugal | 1 (0.6) |  | 1 (2.9) |  |  |  |
| Singapore | 4 (2.3) |  |  |  | 3 (8.6) | 1 (2.9) |
| Spain | 4 (2.3) | 1 (2.9) | 1 (2.9) |  | 1 (2.9) | 1 (2.9) |
| Sweden | 2 (1.1) | 1 (2.9) |  |  |  | 1 (2.9) |
| Switzerland | 1 (0.6) |  |  | 1 (2.9) |  |  |
| Taiwan | 1 (0.6) |  |  | 1 (2.9) |  |  |
| Turkey | 1 (0.6) |  |  |  |  | 1 (2.9) |
| UK | 20 (11.) | 2 (5.7) | 3 (8.6) | 8 (23.) | 4 (11.) | 3 (8.6) |
| USA | 97 (55.) | 25 (71.) | 19 (54.) | 18 (51.) | 19 (54.) | 16 (46.) |

**Appendix Table 7.** Main Study Included Papers Statistics Tools Used, by Epoch.

| Statistics Tools | Overall* | 2010-2013* | 2014-2016* | 2017* | 2018* | 2019* |
| --- | --- | --- | --- | --- | --- | --- |
|  | 175(100) | 35(100) | 35(100) | 35(100) | 35(100) | 35(100) |
| No mention | 28 (16.) | 6 (17.) | 5 (14.) | 8 (23.) | 3 (8.6) | 6 (17.) |
| CART Salford Predictive Miner | 1 (0.6) | 1 (2.9) |  |  |  |  |
| EZR | 1 (0.6) |  |  |  |  | 1 (2.9) |
| Excel | 1 (0.6) |  |  |  |  | 1 (2.9) |
| GraphPad Prism | 1 (0.6) |  |  |  |  | 1 (2.9) |
| JMP | 1 (0.6) | 1 (2.9) |  |  |  |  |
| Mplus | 1 (0.6) | 1 (2.9) |  |  |  |  |
| NCSS | 1 (0.6) |  |  |  |  | 1 (2.9) |
| MedCalc | 1 (0.6) |  | 1 (2.9) |  |  |  |
| Epidata | 1 (0.6) |  |  |  |  | 1 (2.9) |
| R | 20 (11.) |  | 2 (5.7) | 5 (14.) | 9 (26.) | 4 (11.) |
| SAS | 41 (23.) | 8 (23.) | 8 (23.) | 10 (29.) | 10 (29.) | 5 (14.) |
| SigmaPlot | 1 (0.6) |  |  |  |  | 1 (2.9) |
| SPSS (& PASW Statistics) | 50 (29.) | 11 (31.) | 13 (37.) | 10 (29.) | 10 (29.) | 6 (17.) |
| Stata | 37 (21.) | 7 (20.) | 10 (29.) | 3 (8.6) | 8 (23.) | 9 (26.) |
| Statistica | 1 (0.6) | 1 (2.9) |  |  |  |  |
| Statview | 1 (0.6) | 1 (2.9) |  |  |  |  |

**Appendix Table 8.**
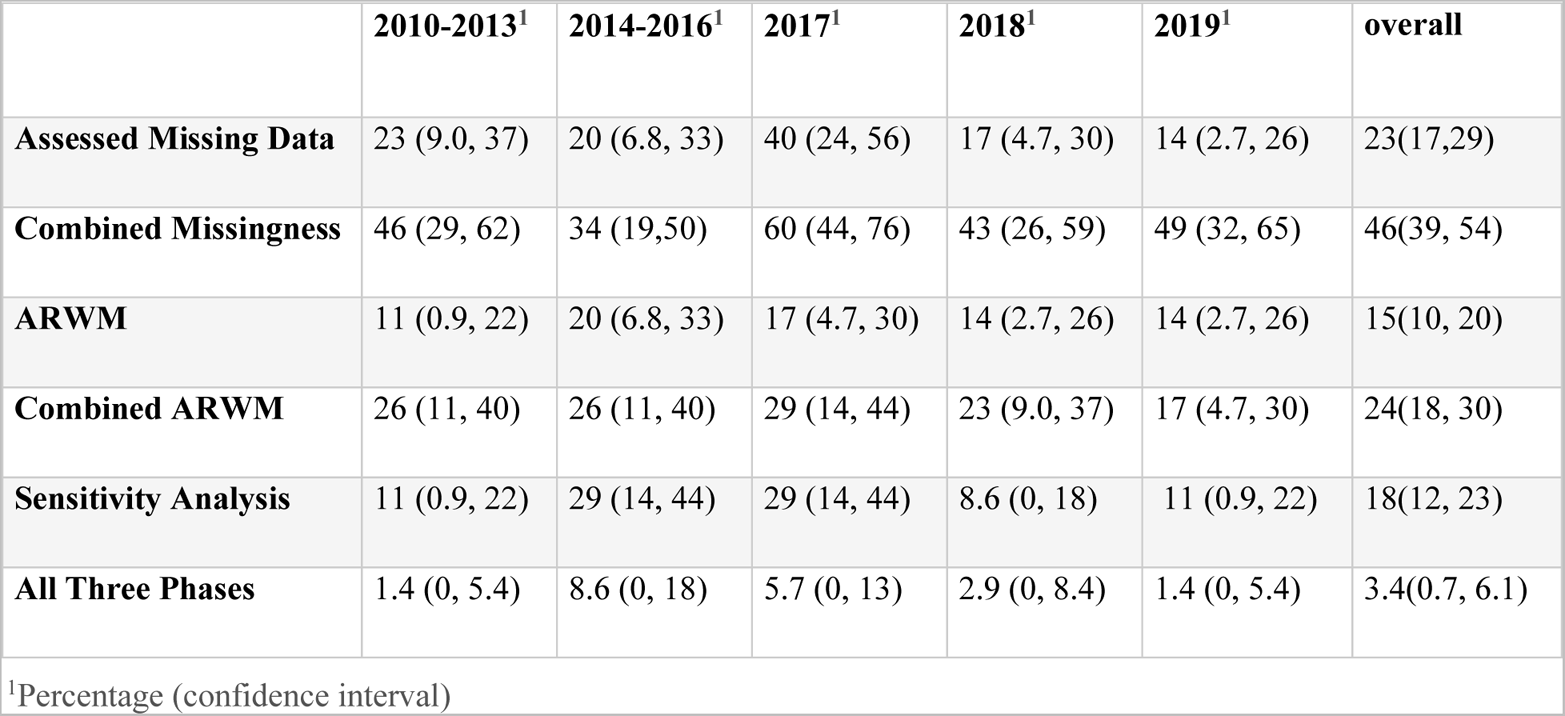
Percentage of Key Methodologies, by Epoch^5^.

|  | 2010-2013<sup>1</sup> | 2014-2016<sup>1</sup> | 2017<sup>1</sup> | 2018<sup>1</sup> | 2019<sup>1</sup> | overall |
| --- | --- | --- | --- | --- | --- | --- |
| **Assessed Missing Data** | 23 (9.0, 37) | 20 (6.8, 33) | 40 (24, 56) | 17 (4.7, 30) | 14 (2.7, 26) | 23(17,29) |
| **Combined Missingness** | 46 (29, 62) | 34 (19,50) | 60 (44, 76) | 43 (26, 59) | 49 (32, 65) | 46(39, 54) |
| **ARWM** | 11 (0.9, 22) | 20 (6.8, 33) | 17 (4.7, 30) | 14 (2.7, 26) | 14 (2.7, 26) | 15(10, 20) |
| **Combined ARWM** | 26 (11, 40) | 26 (11, 40) | 29 (14, 44) | 23 (9.0, 37) | 17 (4.7, 30) | 24(18, 30) |
| **Sensitivity Analysis** | 11 (0.9, 22) | 29 (14, 44) | 29 (14, 44) | 8.6 (0, 18) | 11 (0.9, 22) | 18(12, 23) |
| **All Three Phases** | 1.4 (0, 5.4) | 8.6 (0, 18) | 5.7 (0, 13) | 2.9 (0, 8.4) | 1.4 (0, 5.4) | 3.4(0.7, 6.1) |

<sup>1</sup>Percentage (confidence interval)

**Appendix Table 9.** Article Review Details. Please check https://github.com/ChenyuL/RWE-in-EHR-Data-Analysis/blob/main/ArticleReview_DataCollection.csv

## Footnotes

1 https://github.com/ChenyuL/RWE-in-EHR-Data-Analysis/blob/main/ArticleReview_DataCollection.csv

2 NLM. Epidemiologic Study Characteristics. https://www.ncbi.nlm.nih.gov/mesh/68016020. Published 2020. Accessed 2020/11/09.

3 PubMed. PubMed Clinical Queries https://www.ncbi.nlm.nih.gov/pubmed/clinical. Published 2020. Accessed 2020/11/09, 2020.

4 Detailed annotation and curated data see, https://github.com/ChenyuL/RWE-in-EHR-Data-Analysis

5 Analysis code please see, https://github.com/ChenyuL/RWE-in-EHR-Data-Analysis

